# Study protocol: Strengthening understanding of effective adherence strategies for first-line and second-line antiretroviral therapy (ART) in selected rural and urban communities in South Africa

**DOI:** 10.1101/2021.05.04.21256648

**Authors:** Siphamandla Bonga Gumede, John Benjamin Frank de Wit, Willem Daniel Francois Venter, Samanta Tresha Lalla-Edward

**Author notes:** Corresponding author: Siphamandla Bonga Gumede. **Funding** The principal investigator/SBG is funded by the Utrecht University (UU), Department of Interdisciplinary Social Science, Netherlands (https://www.uu.nl/en/organisation/faculty-of-social-and-behavioural-sciences), and by the Carnegie Corporation of New York (https://www.carnegie.org/) (Grant No--B 8606.R02), Sida (https://www.sida.se/en) (Grant No:54100113), the DELTAS Africa Initiative (https://www.aasciences.africa/aesa/programmes/developing-excellence-leadership-training-and-science-africa-deltas-africa#) (Grant No: 107768/Z/15/Z) and Deutscher Akademischer Austauschdienst (DAAD) (https://www.daad.de/en/). The DELTAS Africa Initiative is an independent funding scheme of the African Academy of Sciences (AAS)–s Alliance for Accelerating Excellence in Science in Africa (AESA) and supported by the New Partnership for Africa–s Development Planning and Coordinating Agency (NEPAD Agency) with funding from the Wellcome Trust (UK) and the UK government. WDFV and STL-E are funded by the National Heart, Lung, And Blood Institute of the National Institutes of Health (https://www.nhlbi.nih.gov/) under Award Number UG3HL156388 and Fogarty International Centre. The content is solely the responsibility of the authors and does not necessarily represent the official views of the National Institutes of Health. The funders had and will not have a role in study design, data collection and analysis, decision to publish, or preparation of the manuscript. **Competing interest** The authors declare that they have no competing interests. **Availability of data and materials Materials** described in this paper are applicable to the study protocol only and there are no raw data reported. The datasets will be collected and can be made available from the corresponding author on reasonable request. Study codebooks, interview guides, informed consent form, participant information sheet, Systematic Review protocol and PRISMA-P checklist developed for this study protocol are included as supporting information (appendix S1, S2, S3, S4, S5, S6). **Consent for publication** Not applicable.

## Abstract

Multiple factors make adherence to antiretroviral therapy (ART) a complex process. This study aims to describe the barriers and facilitators to adherence for patients receiving first-line and second-line ART, identify different adherence strategies utilized and make recommendations for an improved adherence strategy.

This mixed method parallel convergent study will be conducted in seven high volume public health facilities in Gauteng and one in Limpopo province in South Africa. The study consists of four phases; a retrospective secondary data analysis of a large cohort of patients on ART (using TIER.Net, an ART patient and data management system for recording and monitoring patients on ART and tuberculosis (TB) from seven Johannesburg inner-city public health facilities (Gauteng province); a secondary data analysis of the Intensified Treatment Monitoring Accumulation (ITREMA) trial (a randomized control trial which ran from June 2015 to January 2019) conducted at the Ndlovu Medical Center (Limpopo province); in-depth interviews with HIV infected patients on ART (in both urban and rural settings); and a systematic review of the impact of treatment adherence interventions for chronic conditions in sub-Saharan Africa. Data will be collected on demographics, socio-economic status, treatment support, retention in care status, disclosure, stigma, clinical markers (CD4 count and viral load), self-reported adherence information, intrapersonal, and interpersonal factors, community networks, and policy level factors. The systematic review will follow the PRISMA reporting and PICO criteria. Analyses will involve tests of association (Chi-square and t-test), thematic analysis (deductive and inductive approaches) and network meta-analysis.

Using an integrated multilevel socio-ecological framework this study will describe the factors associated with adherence for HIV infected patients who are taking first-line or second-line ART. Implementing evidence-based adherence approaches, when taken up, will improve patient’s overall health outcomes. Our study results will provide guidance regarding context-specific intervention strategies to improve ART adherence.

## Introduction

Inconsistent adherence to treatment is a contributing factor to poor health outcomes of people affected by numerous health conditions, including HIV, tuberculosis, diabetes and hypertension.(1–3). The World Health Organization (WHO) defines adherence as the degree to which a patient is able to follow a treatment schedule and take medication at recommended times (4–6). In the context of HIV, lapses in adherence to medication can lead to the development of viral rebound, which can result in immunosuppression and viral resistance (4,7–9).

For adults and adolescents, WHO recommends tenofovir disoproxil fumarate (TDF) + lamivudine (3TC) (or emtricitabine, FTC) + efavirenz (EFV) as the favoured first-line antiretroviral therapy (ART) combination because of its known safety and efficacy profile (10,11). Despite the advantages of this first-line regimen, in South Africa between 20%-30% of patients with HIV experience clinical, immunological or virological failure from first-line ART due to lapses in adherences (12–14). This is a concern because of the clinical and cost implications attached to treatment failure (15,16).

Adherence to ART is a complex process that is affected by multiple factors, and numerous studies have attempted to establish what the barriers and facilitators of ART adherence are (17–19). Individual-level factors such as age, sex, ethnicity, HIV status disclosure and forgetfulness, have been reported as important in predicting ART adherence (20). However, individual-level factors are only able to report a limited portion of the variability in non-adherence (21). Good interpersonal relationships between patients and care givers or treatment supporters including healthcare providers, an intimate partner, family members, and friends have been reported as a predictor for good adherence (21,22). In contrast to intrapersonal and interpersonal factors, the community level factors such as poverty, HIV related stigma and discrimination against patients on ART introduce barriers to ART adherence (23). Additional to community level factors, healthcare policy level factors like HIV treatment guidelines, policies and best practices are imperative in ensuring good adherence and maintenance of the continuum of care (24).

### Conceptual Framework

Various studies have demonstrated that there are many factors that play an important role in maintaining adherence behaviour (25–28). These factors have been explored using several models, including: 1) Anderson’s Health Care Utilisation model (29), which is a framework that considers predisposing factors (individual’ own personality and behaviour), enabling factors (patient and health provider relationship, community education) and need factors (patient’s beliefs, alternative medicine treatment options, community support); 2) the Dahlgren-Whitehead ‘rainbow model’ (30), a model that builds the relationship between the individual, the environment they live in and health; 3) Information-Motivation-Behavioural skills model (IMB model) (31–33), a model that views adherence behaviour as a function of the interrelations between adherence-related information, motivation, and behavioural skills; 4) the socio-ecological conceptual framework (34,35), which takes into consideration the individual, and their connections to other people, and how they adapt their behaviour to the social environment. This model suggests that an individual’s behaviour is cohesive in a dynamic network of intrapersonal, interpersonal characteristics, community features and existing health policies (34,35).

Our study will adapt the socio-ecological conceptual framework to investigate multilevel and interactive factors such as individual/intrapersonal, interpersonal, community, and health policy level factors that affect adherence to ART (fig 1). It will explore the different aspects of these factors that act either as barriers or facilitators of adherence to ART.

**Fig 1.**
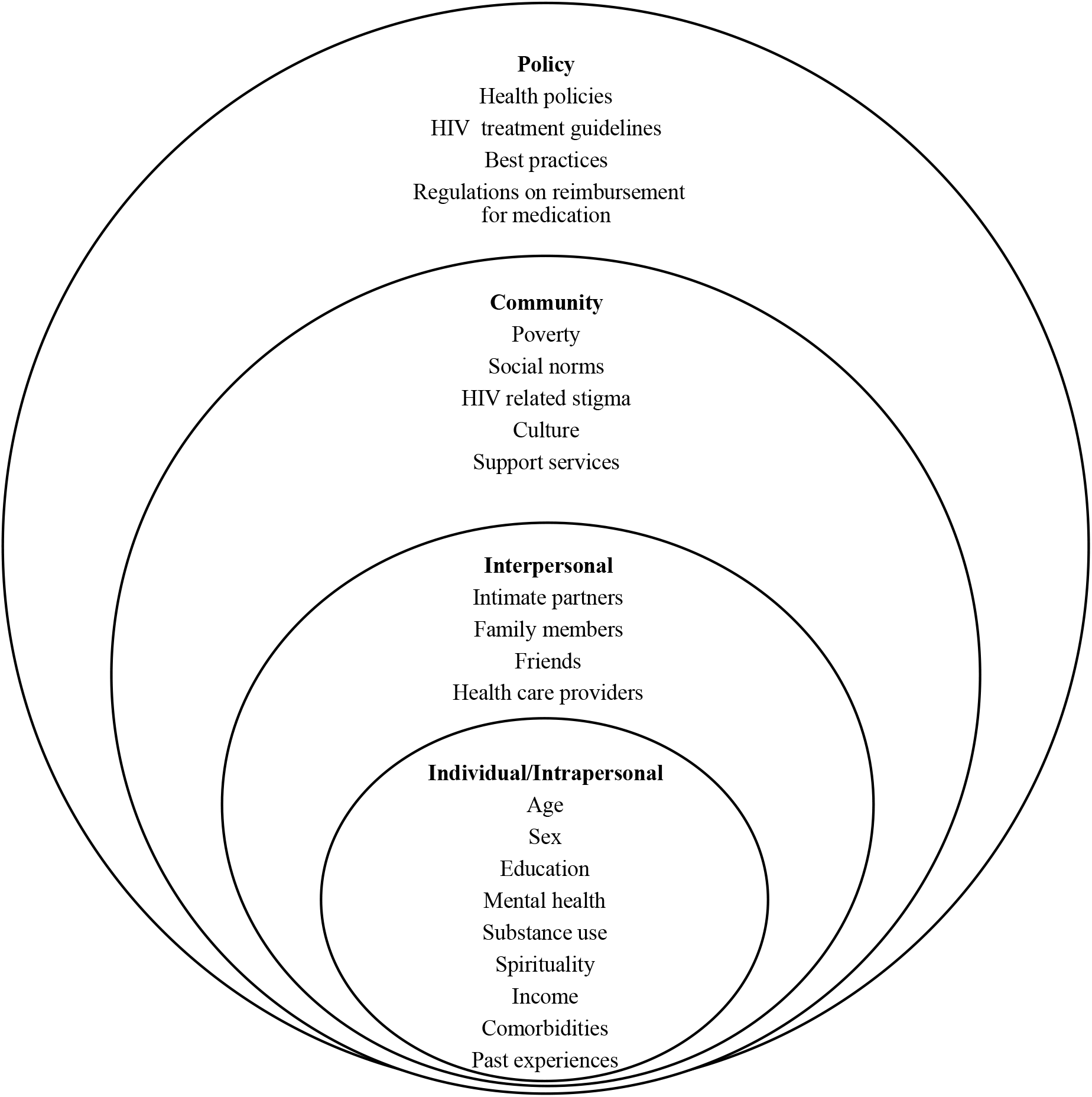
An adapted socio-ecological framework. An adapted socio-ecological framework that depicts the layers of individual, relationship, community, and healthcare policy level factors which influence the processes of treatment adherence and retention in care (35).

#### Intrapersonal level factors

The intrapersonal level of the socio-ecological conceptual framework comprises individual knowledge, attitudes, beliefs, perceptions, and skills that influence behaviour (34). It further includes age, sex, income, mental health, education, substance use, spirituality, comorbidities and past experiences (35).

#### Interpersonal level factors

Here the participant’s social network or relationships with other people including family, friends, peers, intimate partner and health care providers (34) are included. This level also highlights trust and communication as factors that builds a relationship between patients and care givers or treatment supporters (24).

#### Community level factors

The components of the community-level factors that may influence patient’s adherence to ART incorporate cultural views and social norms towards ART access, HIV related stigma, poverty, and available support services within the community (35).

#### Policy level factors

The public policy level is shaped by local, and government laws regarding access and adherence to ART. The socio-ecological conceptual framework reports on macro level factors which includes health policies, HIV treatment guidelines, best practices, and regulations on reimbursement for medication (21,35).

### Research gap

Although ample literature on adherence to ART has been available for a while, there remains a dearth of studies relating to the multi-level factors associated with adherence to treatment and processes shaping adherence behaviour. This lends to the lack of understanding of the interplay between the various factors involved at different levels of the treatment taking behaviour of the ART patient. In South Africa specifically, there is lack of knowledge about the effectiveness of adherence strategies currently employed to promote adherence in healthcare recipients (particularly young patients, males, and those experiencing severe treatment related side effects) and the impact thereof (36,37). Unfortunately, most adherence studies are not guided by any conceptual framework and subsequently are unable to draw adherence strategies from conceptual frameworks of any kind.

### Rationale for the study

Consistent high levels of adherence to HIV medication are important for viral suppression, consequently preventing resistance to ART and progression of the illness (38). According to the socio-ecological framework, numerous levels of factors affects patients’ adherence to treatment (24). Therefore, a multilevel socio-ecological framework will provide information about the influence of particular risk factors relative to others, or if combined effects of risk factors are additive or related. The socio-ecological framework will serve as the conceptual framework in our study in order to understand factors affecting treatment adherence at different levels and also guide strategies to improve ART adherence.

#### Aim

To describe the barriers and facilitators to adherence for patients receiving first-line and second-line ART and different adherence strategies utilised.

### Research questions

1. What are the intra-personal factors (demographic characteristics) and clinical indicators associated with virological failure in first-line and second-line patients in urban communities?
2. What are the intra-and inter-personal factors (demographic, socio-economic characteristics), social and community level factors (poverty, social norms, HIV related stigma, culture), structural factors (health systems, support services) and clinical indicators associated with adherence to first-line and second-line ART in rural communities?
3. What are the perspectives of virally suppressed and unsuppressed first-line and second-line ART patients about treatment adherence in selected urban and rural communities?
  a. Are there any differences between people receiving first-line and second-line ART?
  b. Are there any differences in treatment taking behaviours between virally suppressed and unsuppressed ART patients?
  c. What do virally suppressed and unsuppressed ART patients recommend as adherence strategies?
4. What treatment adherence strategies and interventions for chronic conditions have been tested and implemented in sub-Saharan Africa?

## Materials and Methods

### Study Design

The study will employ a mixed method parallel convergent approach conducted in four phases (I-IV). Due to the different approaches for each study objective, we present the study methods based on each objective.

### Materials and Methods: Phase I

Objective 1: Assess demographic characteristics and clinical indicators associated with virological failure in first-line and second-line patients in an urban community.

#### Study Design

The study will employ a quantitative retrospective cohort study using secondary data analysis of data on people with HIV taking ART (18 years and older) recorded in the TIER.Net database. TIER.Net is the monitoring and evaluation system used by the South African Department of Health for ART patient and data management. It comprises limited demographic information and all treatment and laboratory information from the time of commencement on HIV treatment. This is elaborated on in the data collection section.

The South African Department of Health started providing ART in the public health setting on 01 April 2004. From TIER.Net, we will extract a list of all patients who were initiated on ART from 01 April 2004 to 29 February 2020 in the urban setting (city of Johannesburg region F). The cut-off period of 29 February 2020 was chosen to give ART cohorts a minimum of a 12-month follow-up during which a full clinical assessment could be completed as per guidelines.

#### Study setting

This study will be conducted in seven health facilities in the city of Johannesburg region F. This includes all levels of care:

1. **Primary Health Care**: Jeppe Clinic, Malvern Clinic, Rosettenville Clinic, Yeoville Clinic
2. **Community Health Centre**: Hillbrow Community Health Center (HCHC)
3. **Hospitals**: Charlotte Maxeke Johannesburg Academic Hospital (CMJAH) and South Rand Hospital (SRH)

#### Sampling/Sample Size

For this study, all records of people with HIV who were ever initiated on ART between 01 April 2004 (the inception of the South African national HIV treatment programme in the public health setting) and 29 February 2020 from the seven public health facilities will be included. Based on the TIER.Net database, about 130 000 adult patients were initiated on first-line and second-line ART in the seven facilities selected in the city of Johannesburg region F.

#### Data collection

Data will be extracted from the TIER.Net database (as an MS Excel export file). TIER.Net captures demographic information such as patient age, sex, facility name and contact details and also HIV specific information such as HIV diagnosis date, ART start date, regimen at baseline, ART visit dates, CD4 count, VLD and VLS. The system also records all treatment related information, including ART switch. All these variables will be extracted (see S1 Appendix_study codebook).

The data will be exported to STATA 15.1 for data cleaning and analysis. The control variables are sex, age, level of education and income.

#### Data analysis

Data will be coded and analysed using STATA version 15.1. Tests of association (Chi-square and t-test) between outcome variables and selected socio-demographics and health related characteristics will be conducted. Outcome variables will include (but not limited to) viral load detectability, retention outcomes (active in care, transferred-out, lost to follow-up and dead), side effects and treatment interruptions (stop and restarting treatment). Regression analysis, univariate and multivariate analyses between variables will be built for outcome variables to identify independent predictors. Variables such as age, sex, health facility, beliefs, education, economic status, employment status, religious status, disclosure, and months on ART will be considered as predictor variables or independent variables.

### Materials and Methods: Phase II

Objective 2: Assess demographic, socio-economic characteristics and clinical indicators associated with adherence in patients on first line and second-line ART in a rural community.

#### Study Design

This cohort study using secondary data analysis will be conducted as a sub-study of the Intensified Treatment Monitoring Accumulation (ITREMA) study (39).

The ITREMA study is a randomized control trial that investigated intensified treatment monitoring for HIV patients in rural South Africa (40). The ITREMA database will be used to extract ART information.

#### Study setting

Patient enrolment for the ITREMA trial was done between June 2015 and August 2017 at the Ndlovu Medical Centre in Elandsdoorn, Limpopo Province, South Africa.

#### Sampling/Sample Size

All the records from ITREMA database will be used. There are 501 ART patients in the ITREMA database.

### Data collection

Data will be extracted from the ITREMA databases. The ITREMA database contains fields for demographics, illness, lifestyle, blood tests, alcohol use and medication use. The control variables are sex, age, level of education and income (see S1 Appendix_study codebook). The data will be exported to STATA 15.1 for data cleaning and analysis.

#### Data analysis

Data will be coded and analysed using STATA version 15.1. Tests of association (Chi-square and t-test) between outcome variables and selected socio-demographics and health related characteristics will be conducted. Outcome variables will include (but not limited to) viral load detectability, retention outcomes (active in care, transferred-out, lost to follow-up and dead), side effects and treatment interruptions (stop and restarting treatment). Regression analysis, univariate and multivariate analyses between variables will be built for outcome variables to identify independent predictors. Variables such as age, sex, health facility, beliefs, education, economic status, employment status, religious status, disclosure, and months on ART will be considered as predictor variables or independent variables.

### Materials and Methods: Phase III

Objective 3: Understand adherence in first-line and second-line ART patients who are virologically suppressed and those who are not virologically suppressed in selected urban and rural communities.

#### Study Design

The study will employ a qualitative study design approach.

Active patients from phase I and II (in both urban and rural settings) will be invited to participate in the in-depth interviews (IDIs) to explore factors including (but not limited to) treatment history, current use of ART, treatment regimen, financial/economic factors, risk behaviours (substance use), psychosocial characteristics cultural beliefs, spirituality and), relationship related factors (treatment support), community level factors (societal norms, stigma, discrimination, disclosure), and policy level factors (role of policies and guidelines towards ART adherence).

#### Study setting

This study will be conducted in seven health facilities in the city of Johannesburg region F in Gauteng Province and one in Limpopo Province (Ndlovu Medical Centre).

The seven health facilities in the city of Johannesburg include:

1. **Primary Health Care**: Jeppe Clinic, Malvern Clinic, Rosettenville Clinic, Yeoville Clinic
2. **Community Health Centre**: Hillbrow Community Health Center (HCHC)
3. **Hospitals**: Charlotte Maxeke Johannesburg Academic Hospital (CMJAH) and South Rand Hospital (SRH)

#### Sampling/Sample Size

In this study, purposive sampling of patients currently taking ART will be employed to ensure maximum variation among the study sample. We will ensure a diverse sample by considering viral load status (suppressed and unsuppressed time on ART, age (18 years and older), sex (both males and females) and education. Sample size will depend on when saturation is reached; we anticipate that a maximum of 60 IDIs will be conducted across both study settings and viral load status groups (suppressed and unsuppressed). An anticipated number of study participants to recruit is 15 per viral load status group in each study setting (making a total of 30 participants in each study setting).

#### Data collection

Patients for this phase will be contacted using the contact information that they provided for their facility records or databases used in phase I and II. IDIs will be conducted following a semi-structured interview guide. The guide will comprise open-ended questions covering treatment history, current use of ART and multilevel factors derived from the socio-ecological framework. This will include individual level factors including treatment-related factors (ART regimen, use of non-ART medication), financial and economic factors risk behaviours (substance use), psychosocial factors (cultural beliefs, spirituality), interpersonal-level factors (relationship between patients and treatment supporters or caregivers, such as intimate partners, family members, friends and health care workers), community level factors (social norms regarding HIV and ART, HIV-related stigma and discrimination, HIV-status disclosure), health-system factors (access to HIV and ART services including adherence counselling, and policy level factors (HIV testing and treatment guidelines and policies). Additional probes will be included for each question to promote sharing of detailed information regarding their perspectives and experiences, and to ensure clarification if required (see S1 Appendix_study codebook and S2 Appendix_interview guide). All IDIs will be audio recorded and transcribed verbatim. Transcripts will be translated into English.

#### Data analysis

Transcripts will be imported and analysed using NVIVO. Data coding will be undertaken using deductive (top down) and inductive approaches (bottom up)(41). A deductive approach is driven by researchers’ analytic interest in the study, reflecting the broad issues addressed in the interview guide. An inductive approach is used to identify the detailed themes related to the overarching issues that can be identified in the data (41). Thematic analysis will be used, which is a method for identifying, analysing, and reporting patterns (themes) within data (41,42). Transcripts will be read while noting similar topics that will be grouped into major topics or themes. Data will be analysed as transcripts become available shortly after interviews are conducted. This will ensure the early identification of emerging themes and assist in the identification of data saturation. Analysis of the IDIs will follow the phases of thematic analysis which are familiarization, generating initial codes, searching for themes, reviewing themes and interpretation (41). Familiarization (getting grounded into the data collected), will be achieved by reading the transcripts and field notes repeatedly. During the process of generating codes, key emerging ideas and words from the familiarization phase will be recorded from which we will search, identify, and review themes, concepts, categories, and sub-categories. This will be done in keeping with socio-ecological framework, views and experiences that persist from the data. Finally, factors that influence adherence to ART will be identified and grouped into main categories. Analysis will be guided by the codebook which will be developed by the study team post familiarization with the data. There will be multiple independent coders to ensure the reliability of the coding.

### Materials and Methods: Phase IV

Objective 4: Assess and compare adherence intervention strategies for the chronic conditions of HIV, diabetes mellitus (DM) and hypertension which have been tested and implemented in sub-Saharan Africa (Title: Adherence strategies and interventions for selected chronic conditions in sub-Saharan Africa: a systematic review and meta-analysis) (See S5 Appendix_Systematic review protocol).

#### Study Design

This systematic review will be designed and reported according to the Preferred Reporting Items for Systematic Reviews and Meta-Analysis (PRISMA)(43) (see S6 Appendix_PRISMA checklist), following the registered protocol (CRD42019127564) on the international prospective register of systematic reviews, Prospero (44) (See S5 Appendix_Systematic review protocol). The study will use Population (P), Interventions (I), Comparisons (C) and Outcomes (O) (PICO) criteria as the search strategy tool.

A systematic review on the impact of treatment adherence interventions in chronic conditions (HIV, hypertension, diabetes mellitus) in sub-Saharan Africa will be conducted to provide context to adherence in sub-Saharan Africa. In this region, HIV remains the leading cause of death more especially in the young and middle-aged adults. However, the burden of non-communicable diseases (NCDs), particularly DM and hypertension, has increased rapidly in recent years (45–47).

#### Study setting

All information from sub-Saharan Africa only will be included for the systematic review.

#### Sampling/Sample Size

All interventions described as chronic conditions adherence interventions (HIV/ART, hypertension, DM). Inclusion in the systematic review will be dependent on the criteria set out in the systematic review protocol (44) and reported using the PRISMA reporting guidelines.

#### Data collection

A pre-defined data sheet will be developed for data extraction. The tool will include (but not be limited to): reference (author, title), year of publication, setting or location, sample size, intervention description, participants receiving adherence (in case of comparison) (see S1 Appendix_study codebook). The form/tool will be tested before conducting the final searches. One reviewer will conduct all the data extraction while a second reviewer will be responsible for data quality assurance on the extraction and also conduct full text review of the included material. We will search using several electronic databases. These will include PubMed/Medline, Web of Science, Google Scholar, Scopus, and CINAHL. If necessary, we will contact study authors and request more information on individual studies. Citations and bibliographies of records will be reviewed to identify additional relevant material.

The basic search terms included will be:

*‘Chronic conditions’* OR *‘hypertension’* OR *‘high blood pressure’* OR *‘blood pressure’* OR *‘arterial hypertension’* OR *‘mellitus diabetes type I’* OR *‘mellitus diabetes type II’* OR *‘Diabetes’* OR *‘Sugar’* OR *‘HIV’* OR *‘Antiretroviral Therapy’* OR *‘Antiretroviral Treatment’* OR *‘ART’* OR *‘ART Programs’* OR *‘ART Programmes’* AND *‘adherence’* OR *‘compliance’* AND *‘interventions’* OR *‘strategies’* OR *‘odds ratio’* OR *‘risk ratio’* OR *‘evaluation’* OR *‘impact’* OR *‘effectiveness’* OR *‘outcome’* AND *‘sub-Saharan Africa’* OR *‘sub Saharan Africa’* OR *‘sub-Saharan African’* OR *‘sub Saharan African’* OR *‘Africa’ (table 1)*.

**Table 1:**
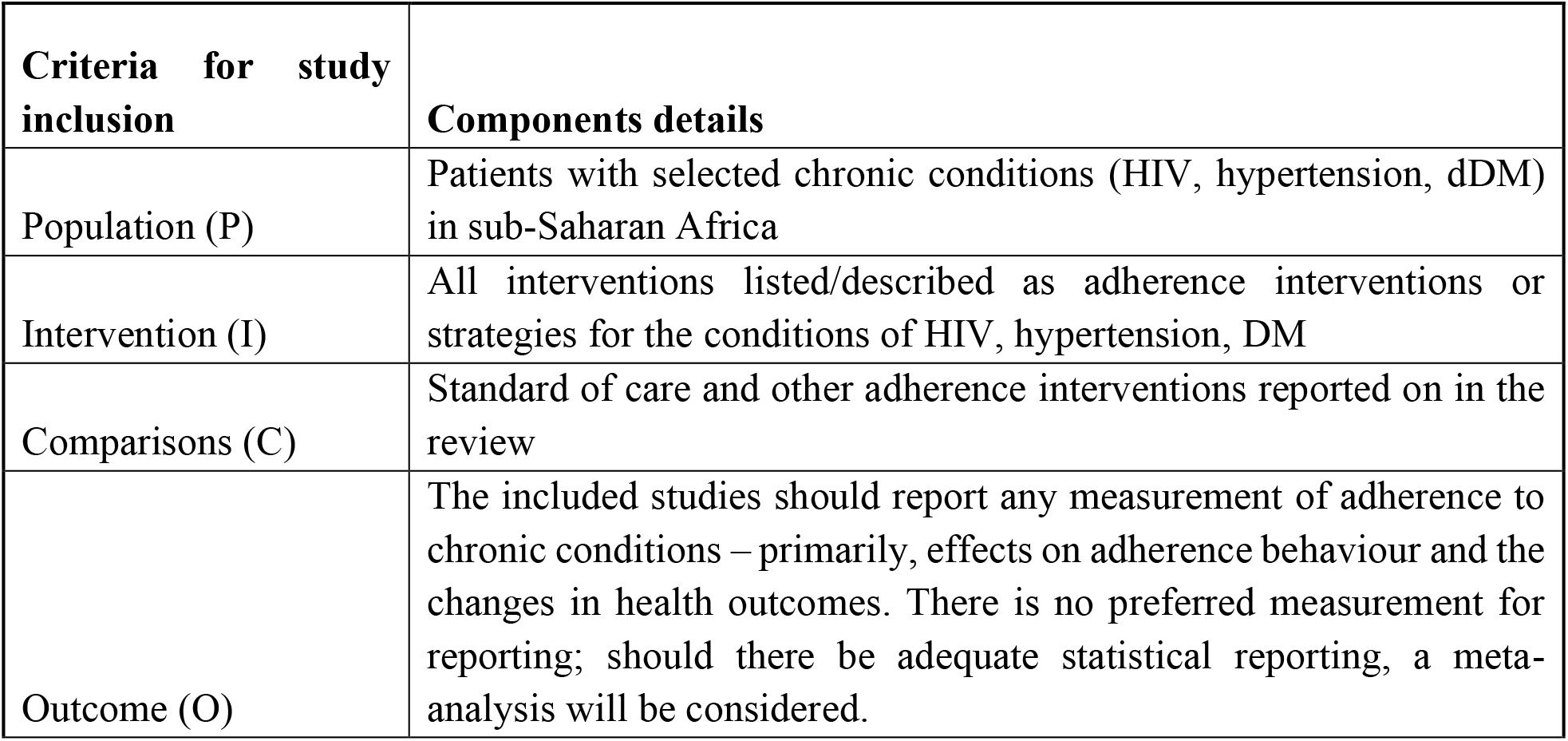
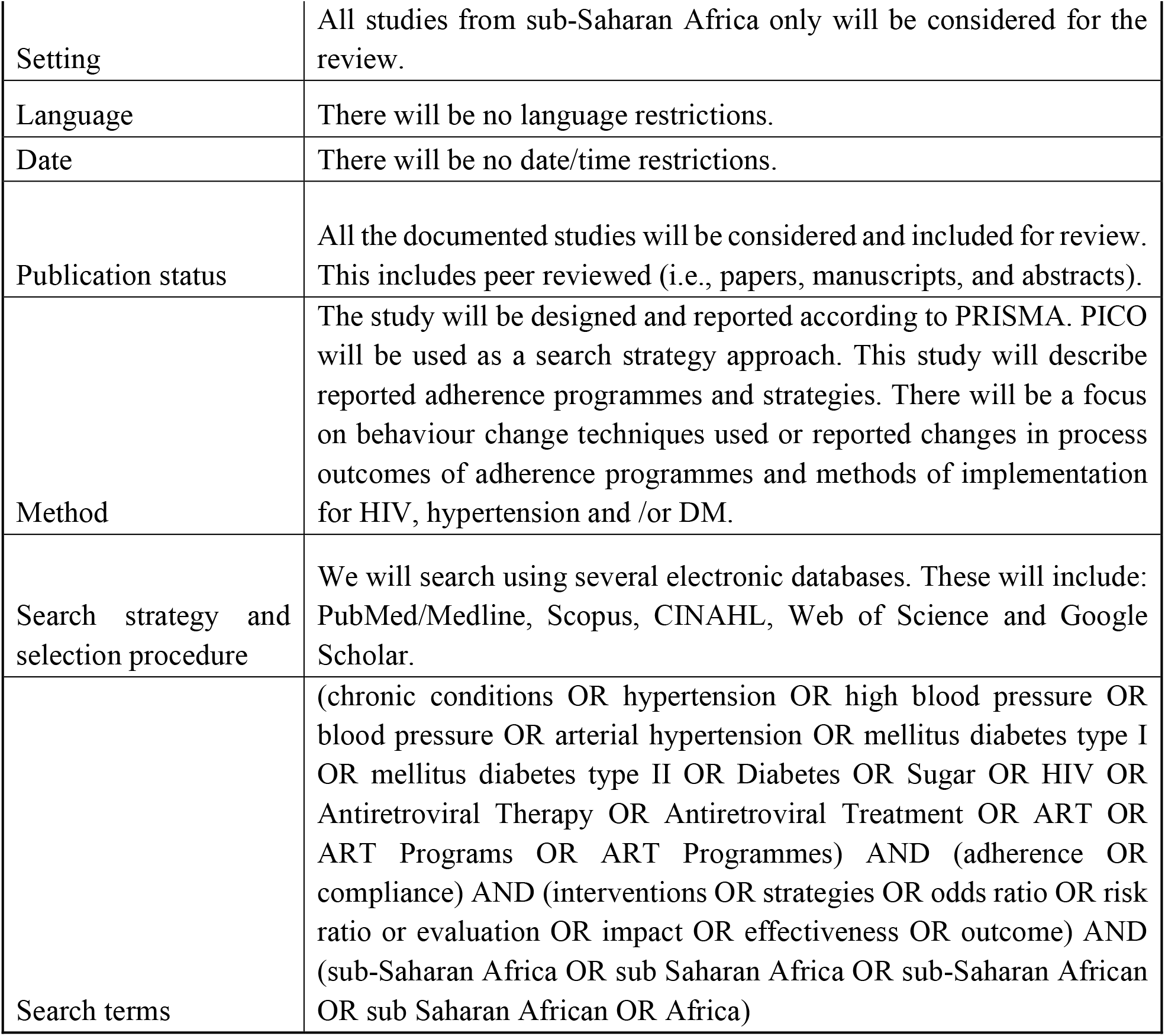
Methodological aspect of the systematic review.

| Criteria for study inclusion | Components details |
| --- | --- |
| Population (P) | Patients with selected chronic conditions (HIV, hypertension, dDM) in sub-Saharan Africa |
| Intervention (I) | All interventions listed/described as adherence interventions or strategies for the conditions of HIV, hypertension, DM |
| Comparisons (C) | Standard of care and other adherence interventions reported on in the review |
| Outcome (O) | The included studies should report any measurement of adherence to chronic conditions – primarily, effects on adherence behaviour and the changes in health outcomes. There is no preferred measurement for reporting; should there be adequate statistical reporting, a meta-analysis will be considered. |
| Setting | All studies from sub-Saharan Africa only will be considered for the review. |
| Language | There will be no language restrictions. |
| Date | There will be no date/time restrictions. |
| Publication status | All the documented studies will be considered and included for review. This includes peer reviewed (i.e., papers, manuscripts, and abstracts). |
| Method | The study will be designed and reported according to PRISMA. PICO will be used as a search strategy approach. This study will describe reported adherence programmes and strategies. There will be a focus on behaviour change techniques used or reported changes in process outcomes of adherence programmes and methods of implementation for HIV, hypertension and /or DM. |
| Search strategy and selection procedure | We will search using several electronic databases. These will include: PubMed/Medline, Scopus, CINAHL, Web of Science and Google Scholar. |
| Search terms | (chronic conditions OR hypertension OR high blood pressure OR blood pressure OR arterial hypertension OR mellitus diabetes type I OR mellitus diabetes type II OR Diabetes OR Sugar OR HIV OR Antiretroviral Therapy OR Antiretroviral Treatment OR ART OR ART Programs OR ART Programmes) AND (adherence OR compliance) AND (interventions OR strategies OR odds ratio OR risk ratio or evaluation OR impact OR effectiveness OR outcome) AND (sub-Saharan Africa OR sub Saharan Africa OR sub-Saharan African OR sub Saharan African OR Africa) |

The search terms will be adjusted to suit the database being searched. An inventory with the database searched, the corresponding search criteria used, the date when the searches were conducted, and the results will be maintained. A second reviewer will run the searches separately for comparison. The strength of the body of evidence (quality of evidence), the risk of bias and magnitude of effect will be rated and assessed using Grading of Recommendations Assessment, Development and Evaluation (GRADE) (48,49).

#### Data analysis

All adherence interventions or strategies will be described, based on the type of intervention implemented and the setting. The different evaluations methods will then be described in detail by comparing the type of assessments and outcome measures. If appropriate, outcome measures will be reported in terms of changes in the prevalence or reduction in the relative risk. Whenever necessary, we will calculate unadjusted risk ratios (RRs) and 95% confidence intervals (CIs) from data provided and present the outcome indicator results in forest plots. Furthermore, we will perform a sensitivity analysis to measure the robustness of our results to the choice of summary statistic and calculated unadjusted risk differences. We will apply a random-effects model to calculate summary RRs and 95% Cl. To test the robustness of the findings, we will re-run the analysis using a fixed effects model. Data will be coded and analysed used STATA version 15.1.

Details for study criteria are presented in table 1

### Data management/Data cleaning (phase I-IV)

Data quality scripts for phase I, II, IV will be written in STATA. Data quality checks for phases I, II, and IV (quantitative data) will be done through RedCap (a secure web platform for building and managing research databases). Since TIER.Net will be used as the primary data source for phase I data extraction in the city of Johannesburg region F, where there are data quality issues, the study team will liaise with the facility staff and developmental partners in the region to assist with data clean-up activities. Data quality issues with the ITREMA data (phase II) will be communicated to the ITREMA study quality assurance officer for rectifying. Data quality checks for phase III (qualitative data) will be done in Microsoft Word (before exporting data to NVIVO). All transcripts will be checked for completeness and accuracy against original interviews. Creditability of data analysis will be ensured through triangulation of data sources (i.e., original interviews, field notes, transcripts, medical record).

### Data storage and access (phase I-IV)

Data will be captured and stored electronically, and password protected in the Microsoft Word, Microsoft Excel format and/or RedCap and will only be accessible to an investigator and supervisors only. RedCap access is restricted to only those users who are registered on the system. All data from RedCap, Microsoft Word, Microsoft Excel, STATA, and voice files will be stored in access restricted folders on the Ezintsha server which will only be accessible to an investigator and supervisors. Any paper versions of data will be discarded after use. Data storage and access measures will also comply with data storage and access requirements of the Utrecht University.

### Ethical approvals and consent to participate

We obtained ethical clearance from the University of the Witwatersrand Human Research Ethics Committee (clearance certificate number: M190641). Departmental approval was granted by the Johannesburg Health District (DRC Ref: 2019-10-005 and National Health Research Database reference number: GP_201910_031). Written consent for interviews will be obtained from all participants (see S3 Appendix_informed consent form). All participants will be provided with written information about the research, and they will also be verbally informed that their participation is voluntary and that they may withdraw from participation at any time (see S4 Appendix_Information sheet).

## Discussion

The overarching aim of this study is to contribute to knowledge that can provide guidance regarding the barriers and facilitators to adherence for first-line and second-line ART patients and different adherence strategies utilized. Using an integrated multilevel socio-ecological framework, this study will focus on determining the influence of the multiple factors that impact on adherence to ART. In line with socio-ecological frameworks (50,51) and propositions, the findings from this study will be discussed under four units of analysis: intrapersonal level, interpersonal level, community level and policy level factors.

### Intrapersonal factors

The client knowledge, attitudes, experiences and perceptions coupled with analysis of the intrapersonal factors influencing adherence to ART play a fundamental role in maintaining adherence (34). Intrapersonal level factors such as being age, sex, substance abuse, and comorbidities will be discussed, in line with the existing literature (52–55).

### Interpersonal level factors

The trust and communication between patients on ART and treatment supporters is essential in improving and maintaining optimal adherence to treatment (24). Some studies have reported treatment support as a predictor of adherence (37,56). In this study, the interpersonal level factors consisting of relationships between patients and family members, friends, intimate partner(s) and healthcare providers will be discussed against previous studies to provide guidance on the role of treatment supporters in strengthening treatment adherence.

### Community level factors

The findings concerning community level factors will consider the importance of patients understanding of social norms, cultural barriers and reduction of poverty, stigma and discrimination against people living with HIV and on ART (57). Information on poverty, culture, HIV related stigma, and discrimination caused by misconceptions will be discussed in line with existing literature to provide a critical assessment on the role of community level factors on ART adherence.

### Policy level factors

The policy-level factors address knowledge and influence of public health policies, guidelines, and standards to patients (58,59). The South African HIV programme has undergone several changes since its implementation in 2004 (60). This study’s findings will demonstrate the participant’s understanding of existing ART adherence or HIV treatment related health policies and guidelines and best practices that are followed by health providers when providing health services to patients. Knowledge of ART medicines or regimens (names of ART drugs), definitions, and clinical functions of viral load (knowledge of threshold for virological failure and suppression) and CD4 cell count (understanding of high or low CD4 cell count) will be discussed as themes in assessing individual’s knowledge of existing HIV treatment policies and guidelines. Additionally, we will be able to provide recommendations on required changes at the policy level to improve treatment adherence.

In conclusion, our study will demonstrate how an existing socio-ecological conceptual framework can be used as a tool to provide guidance regarding facilitators and barriers to ART adherence. By populating this framework through secondary data analysis, participant interviews of HIV infected people who are taking ART and a systematic review comparing adherence intervention strategies for the chronic conditions, this mixed method study will provide evidence on factors affecting treatment adherence at different socio-ecological levels and guide context-specific intervention strategies to improve ART adherence. We believe that the use of our study results to strengthen adherence intervention will subsequently improve health outcomes and decrease the number of patients switching to complex treatment such as second-line and third-line regimens.

### Dissemination

Findings from this research will be submitted for doctoral degree purposes (by thesis). Peer reviewed publications and scientific conference presentations will be developed. Results of the research will be shared with the research participants, donors, and health facilities.

## Supporting information

FUNDING AGREEMENT CARNIEGE AND SIDA

FUNDING AGREEMENT UTRECHT UNIVERSITY

Human right research ethics clearance letter

Department of health letter of permission

Study approval from CMJA hospital (departmental approval)

Study approval from South Rand hospital (departmental approval)

Wits Faculty of Health Science approval of study title

## Data Availability

Materials described in this paper are applicable to the study protocol only and there are no raw data reported. The datasets will be collected and can be made available from the corresponding author on reasonable request. Study codebooks, interview guides, informed consent form, participant information sheet, Systematic Review protocol and PRISMA-P checklist developed for this study protocol are included as supporting information (appendix S1, S2, S3, S4, S5, S6).

## Authors’ contributions

**Conceptualization:** Siphamandla Bonga Gumede, Samanta Tresha Lalla-Edward, Willem Daniel Francois Venter, John Benjamin Frank de Wit

**Funding acquisition:** Siphamandla Bonga Gumede, Samanta Tresha Lalla-Edward, Willem Daniel Francois Venter, John Benjamin Frank de Wit

**Supervision:** Samanta Tresha Lalla-Edward, Willem Daniel Francois Venter, John Benjamin Frank de Wit

**Methodology:** Siphamandla Bonga Gumede, Samanta Tresha Lalla-Edward, Willem Daniel Francois Venter, John Benjamin Frank de Wit

**Writing – Original Draft Preparation:** Siphamandla Bonga Gumede, Samanta Tresha Lalla-Edward

**Writing – Review & Editing:** Siphamandla Bonga Gumede, Samanta Tresha Lalla-Edward, Willem Daniel Francois Venter, John Benjamin Frank de Wit

## Acknowledgments

This research is supported by the Consortium for Advanced Research Training in Africa (CARTA). CARTA is jointly led by the African Population and Health Research Center and the University of the Witwatersrand. The statements and views made in this article are solely the responsibility of the authors.

We would like to thank all the relevant health and research authorities from the City of Johannesburg and Ndlovu Medical Center for allowing the research team to engage in a partnership to strengthen health service delivery through technical assistance and research.

## Notes

### Competing Interest Statement

The authors have declared no competing interest.

### Author Declarations

We obtained ethical clearance from the University of the Witwatersrand Human Research Ethics Committee (clearance certificate number: M190641). Departmental approval was granted by the Johannesburg Health District (DRC Ref: 2019-10-005 and National Health Research Database reference number: GP_201910_031). Written consent for interviews will be obtained from all participants.

