## Supplementary appendix 1 for "Study protocol: Strengthening understanding of effective adherence strategies for first-line and second-line antiretroviral therapy (ART) in selected rural and urban communities in South Africa"

**CODEBOOK FOR ALL STUDY PHASES**

**Phase I codebook: Secondary data analysis of a large cohort of patients on ART (from TIER.Net database)**

| **Variables (from TIER.Net)** | **Description and codes** |
|---|---|
| Patient unique number | Unique identity of a patient. The unique number vary by facilities and may also assist in picking duplicates |
| Facility | Name of a corresponding facility |
| Name | Name of the client/patient |
| Surname | Surname of the client/patient |
| Date Of Birth | The date of birth of the client/patient |
| Gender | Male of Female |
| Age At Pre-ART | This is applicable to patients who attended pre-Art prior to ART initiation and may be seen in older patients (before Universal Test and Treat) |
| Age At ART Start | This reflects age of the patient when started ART or restarted ART in case of defaulters or LTFU |
| Current Age | Refers to age of the patient at present (18 years and older) |
| First Visit Date | The first date the patient was seen in the facility (This date may be the same date as the diagnosis date) |
| HIV Diagnosis Date | Date the patient was confirmed as HIV positive |
| ART Start Date | Date of the patient’s first ever experience with triple therapy (date when patient was started on ART for the first time ever) or re-initiated in case of Exp patients |
| Baseline CD4 | First ever CD4 count done after HIV positive diagnosis (sometimes before starting the patient on ART). Before 2016, CD4 count was done after HIV positive status to assess patient's ART elgibility |

| | |
|---|---|
| Regimen At Baseline | First ever regimen issued to the patient when initiated on ART for the first time or when restated on ART (in case of defaulters or LTFU) |
| Method into ART | Patient has started ART at one facility and has now transferred to another facility |
| Method Into ART Location | For Transferred/Moved-In patients, facility/location where the patients comes from (referring facility) |
| Transferred/Moved In Date | If client is a Transferred/Moved-in, Date when the client was transferred in the current clinic |
| Outcome | "Outcome" reflects active or inactive status of patients (whether the patient is still attending ART services in the facility or not). For active patients, this variable will be blank (however few blanks may also reflects unconfirmed lost to follow-up and this should be verified against Last ART visit date of the patient. In active patients will be coded as either Lost-to-follow up (LTFU), Transferred/Moved out and Died |
| Outcome Date | For inactive patients (LTFU, Transferred/Moved Out, Died), outcome date reflects date outcome was confirmed. For LTFU and Transferred/Moved Out-it reflects the last time the patient was at the facility |
| Prior ART | Reflects whether patient has ever taken ART before ART start date. Options listed as NO ART EXP (naïve), PMTCT, Prior ART > 30 days (EXPERIENCE), |
| Last ART Visit Code | Regimen that was given/issued to the patient at a last visit |
| Last ART Visit Date | The last visit date when the patient was seen in the facility. It is the date when the Last ART visit code was provided |
| Last ART CD4 Count | The last or latest CD4 count/result captured on TIER.Net. It is important to highlight that this depends on capturing and data quality i.e. blood results capturing into TIER.Net |
| Last ART CD4 Date | The date of the Last CD4 Count captured on TIER.Net |
| Last ART Next Appointment Date | This refers to the next ART appointment given to the patient at last ART visit. This date assists in ensuring that LTFU is calculated accurately |
| TB Rx Started | Did the client start on TB treatment (Yes,No, Not Sure). This is not at |

| | |
|---|---|
| Last ART VL Count | The last or latest Viral Load result captured on TIER.Net. It is important to highlight that this depends on capturing and data quality i.e. blood results capturing into TIER.Net |
| Last ART VL Date | The date of the Last Viral Load results captured on TIER.Net. |
| Second Line Start Date | Second Line Start Date. This is the date a patient was switched from first line to a second line regimen |
| Last ART Cell Code | Refers to the last regimen issued to the patient or last regimen that was expected to be issued to the patient. This includes the expected visits or appointments where the patients were unable to attend the clinic ("Did Not Attend"). |
| Last ART Cell Date | Refers to the date of the Last ART Cell Code. With the description of a Last ART Cell Code, this might therefore be a future date when looked at against Last ART Visit Date |
| Last ART Prescription Code | Refers to regimen prescribed to the patient at a last ART visit. Sometimes this is similar to Last ART Visit Code |
| Last ART Prescription Date | This reflects the date of the last prescription provided to the patient. This may be similar to Last ART Visit Date |
| TB Status At Last Visit | Reflects TB screening and dianosis at last visit. This includes the following options No Symptoms, Symptoms-With Sputum, Symptoms-No Sputum, On TB Treatment at this facility, On TB treatment at another facility, Not Screened, Screening Status Unknown |
| Pregnant on ART Start | Was the client/patient pregnant when starting ART |
| Pap Smear | Reflects whether pap smear was done to the patient (female) |
| Pap Smear Date | Reflects pap smear test date, if pap smear was done |
| Last Sub Clinic | Name of the last Sub Clinic. This includes adherence clubs which mostly second line patients will not be allocated to and in case this is captured, data quality may be required |
| Health Provider At ART Start | Name of the health provider who initiated/started the patient on ART |
| Pregnancy Status At Last Visit | Refers to pregnancy status in female patients at a last visit (listed as Yes, No or Not Sure). Screening for pregnancy is done at all visits unless the patient is pregnant |

| | |
|---|---|
| ART Restart Date After LTFU | In case the patient defaulted or was declared a LTFU, a date the patient was restarted on ART |
| Duration On ART (Months) | A period the patient has been on ART since first initiation or re-initation (for experienced patients) |

**Phase II codebook: Secondary data analysis for the cohort from the ITREMA clinical trial**

| Question | Code | Numeric or textual | 0 | 1 | 2 | 3 | 4 | 5 | 6 | 7 | 8 | 9 |
|---|---|---|---|---|---|---|---|---|---|---|---|---|
| id | entry_no | 1-600 | | | | | | | | | | |
| part_id | participant_no | ITREMA001ITREMA600 | | | | | | | | | | |
| q4 | employment | | _ | | y_self | n_unempl | n_stud | n_ret | n_dis | n_oth | ref | dk |
| q5 | personal_pay | 0-100000 | | | | | | | | | | |
| q6a | hh_in_salaries | 0-100000 | | | | | | | | | | |
| q6b | hh_in_business | 0-100000 | | | | | | | | | | |
| q6c | hh_in_remittance | 0-100000 | | | | | | | | | | |
| q6d | hh_in_pension | 0-100000 | | | | | | | | | | |
| q6e | hh_in_grants | 0-100000 | | | | | | | | | | |
| q6f | hh_in_other | 0-100000 | | | | | | | | | | |
| q7a | grant_age | | no | yes | | | | | | | | |
| q7b | grant_disability | | no | yes | | | | | | | | |
| q7c | grant_child_sup | | no | yes | | | | | | | | |
| q7d | grant_care | | no | yes | | | | | | | | |
| q7e | grant_foster_child | | no | yes | | | | | | | | |

| | | | | | | | |
|---|---|---|---|---|---|---|---|
| q7f | grant_veteran | | no | yes | | | |
| q7g | grant_in_aid | | no | yes | | | |
| q7h | grant_social | | no | yes | | | |
| q7i | grant_refused | | no | yes | | | |
| q7j | NA | NA | NA | NA | NA | NA | NA |

| q8a | rela_status | | _ | married | life_par t | half | single | divorc ed | widow ed | multi ple | oth er |
|---|---|---|---|---|---|---|---|---|---|---|---|
| q8b | rela_start | date | | | | | | | | | |
| q8c | rela_other | text | | | | | | | | | |
| q9 | people_under_roo f | 0-20 | | | | | | | | | |
| q10a | hh_partner | 0-5 | | | | | | | | | |
| q10b | hh_child_bio | 0-20 | | | | | | | | | |
| q10c | hh_child_other | 0-20 | | | | | | | | | |
| q10d | hh_parent | 0-2 | | | | | | | | | |
| q10e | hh_in law | 0-2 | | | | | | | | | |
| q10f | hh_sibling | 0-20 | | | | | | | | | |
| q10g | hh_grandparent | 0-8 | | | | | | | | | |
| q10h | hh_other_fam | 0-20 | | | | | | | | | |

| | | | | | | |
|---|---|---|---|---|---|---|
| q10i | hh_non_relative | 0-20 | | | | |
| q10j | hh_refused | | no | yes | | |
| q10k | hh_dk | | no | yes | | |
| q11 | no_money_food | | _ | yes | no | dk |
| q12 | nf_recent | | n/a | yes | no | dk |
| q13 | nf_how_often | | n/a | yes | no | dk |
| q14 | nf_hungry | | n/a | yes | no | dk |
| q15a | nf_jan | | no_n/a | yes | | |
| q15b | nf_feb | | no_n/a | yes | | |
| q15c | nf_mar | | no_n/a | yes | | |
| q15d | nf_apr | | no_n/a | yes | | |
| q15e | nf_may | | no_n/a | yes | | |
| q15f | nf_jun | | no_n/a | yes | | |
| q15g | nf_jul | | no_n/a | yes | | |
| q15h | nf_aug | | no_n/a | yes | | |

| | | | | | | | | |
|---|---|---|---|---|---|---|---|---|
| q15i | nf_sep | | no_n/a | yes | | | | |
| q15j | nf_oct | | no_n/a | yes | | | | |
| q15k | nf_nov | | no_n/a | yes | | | | |
| q15l | nf_dec | | no_n/a | yes | | | | |
| q15m | nf_dk | | no_n/a | yes | | | | |
| q16 | ARV_ability | | _ | compl_unsure | unsure | sure | compl_sure | |
| q17 | ARV_positive_effect | | _ | compl_unsure | unsure | sure | compl_sure | |
| q18 | ARV_resistance | | _ | compl_unsure | unsure | sure | compl_sure | |
| q19 | ss_satisfaction | | _ | v_dissat | sw_dissat | sw_satis | v_satis | |
| q20 | ss_remember_medication | | _ | not_at_all | somewhat | a_little | a_lot | n/a |
| q21 | md_away | | _ | never | rarely | sometimes | often | n/a |
| q22 | md_busy | | _ | never | rarely | sometimes | often | n/a |
| q23 | md_forgot | | _ | never | rarely | sometimes | often | n/a |
| q24 | md_too_many | | _ | never | rarely | sometimes | often | n/a |
| q25 | md_side_effect | | _ | never | rarely | sometimes | often | n/a |

| | | | | | | | | | | |
|---|---|---|---|---|---|---|---|---|---|---|
| q26 | md_privacy | | _ | never | rarely | sometimes | often | n/a | | |
| q27 | md_routine | | _ | never | rarely | sometimes | often | n/a | | |
| q28 | md_harmful | | _ | never | rarely | sometimes | often | n/a | | |
| q29 | md_asleep | | _ | never | rarely | sometimes | often | n/a | | |
| q30 | md_sick | | _ | never | rarely | sometimes | often | n/a | | |
| q31 | md_depression | | _ | never | rarely | sometimes | often | n/a | | |
| q32 | md_specific_time | | _ | never | rarely | sometimes | often | n/a | | |
| q33 | md_empty | | _ | never | rarely | sometimes | often | n/a | | |
| q34 | md_felt_good | | _ | never | rarely | sometimes | often | n/a | | |
| q35 | often_on_time | | _ | all_time | most_time | rarely | never | n/a | | |
| q36 | md_days_week | | _ | every_day | 4_to_6 | 2_to_4 | once | less_once | never | n/a |
| q37 | md_last time | | _ | past_week | 1_to_2w | 2_to_4w | 1_to_3m | more_3m | never | n/a |
| q38 | hh_supp_work | | _ | no_sup | little_sup | fair_sup | lot_sup | | | |
| q39 | hh_supp_worries | | _ | no_sup | little_sup | fair_sup | lot_sup | | | |
| q40 | hh_supp_leisure | | _ | no_sup | little_sup | fair_sup | lot_sup | | | |

| | | | | | | | | |
|---|---|---|---|---|---|---|---|---|
| q41 | hh_supp_practical | | _ | no_sup | little_s up | fair_su p | lot_sup | |
| q42 | hh_supp_personal | | _ | no_sup | little_s up | fair_su p | lot_sup | |
| q43 | non_hh_supp_wor k | | _ | no_sup | little_s up | fair_su p | lot_sup | |
| q44 | non_hh_supp_wor ries | | _ | no_sup | little_s up | fair_su p | lot_sup | |
| q45 | non_hh_supp_leis ure | | _ | no_sup | little_s up | fair_su p | lot_sup | |
| q46 | non_hh_supp_pra ctical | | _ | no_sup | little_s up | fair_su p | lot_sup | |
| q47 | non_hh_supp_per sonal | | _ | no_sup | little_s up | fair_su p | lot_sup | |
| q48 | stress_get_away | | _ | never | rarely | someti mes | v_often | always |
| q49 | stress_solve_probl em | | _ | never | rarely | someti mes | v_often | always |
| q50 | stress_blame_situ ation | | _ | never | rarely | someti mes | v_often | always |
| q51 | stress_treat_food | | _ | never | rarely | someti mes | v_often | always |
| q52 | stress_anxious | | _ | never | rarely | someti mes | v_often | always |
| q53 | stress_similar_pro blem | | _ | never | rarely | someti mes | v_often | always |
| q54 | stress_visit_friend | | _ | never | rarely | someti mes | v_often | always |
| q55 | stress_determine_ action | | _ | never | rarely | someti mes | v_often | always |

| | | | | | | | | | |
|---|---|---|---|---|---|---|---|---|---|
| q56 | stress_buy_me | | _ | never | rarely | someti mes | v_often | always | |
| q57 | stress_blame_emo tional | | _ | never | rarely | someti mes | v_often | always | |
| q58 | stress_work_unde rstand | | _ | never | rarely | someti mes | v_often | always | |
| q59 | stress_upset | | _ | never | rarely | someti mes | v_often | always | |
| q60 | stress_corrective_ action | | _ | never | rarely | someti mes | v_often | always | |
| q61 | stress_blame_kno wing | | _ | never | rarely | someti mes | v_often | always | |
| q62 | stress_special_per son | | _ | never | rarely | someti mes | v_often | always | |
| q63 | stress_think_learn | | _ | never | rarely | someti mes | v_often | always | |
| q64 | stress_wish_chang e | | _ | never | rarely | someti mes | v_often | always | |
| q65 | stress_go_food | | _ | never | rarely | someti mes | v_often | always | |
| q66 | stress_analyze_rea ct | | _ | never | rarely | someti mes | v_often | always | |
| q67 | stress_inadequaci es | | _ | never | rarely | someti mes | v_often | always | |
| q68 | stress_phone_frie nd | | _ | never | rarely | someti mes | v_often | always | |
| q69 | hcw_depend | | _ | stro_disag r | disagr | sli_disa gr | sli_agre e | agree | stro_ag ree |
| q70 | hcw_understand | | _ | stro_disag r | disagr | sli_disa gr | sli_agre e | agree | stro_ag ree |

| | | | | | | | | | |
|---|---|---|---|---|---|---|---|---|---|
| q71 | hcw_distrust | | _ | stro_disagr | disagr | sli_disagr | sli_agree | agree | stro_agree |
| q72 | hcw_joint_effort | | _ | stro_disagr | disagr | sli_disagr | sli_agree | agree | stro_agree |
| q73 | hcw_similar_ideas | | _ | stro_disagr | disagr | sli_disagr | sli_agree | agree | stro_agree |
| q74 | hcw_respect | | _ | stro_disagr | disagr | sli_disagr | sli_agree | agree | stro_agree |
| q75 | hcw_like | | _ | stro_disagr | disagr | sli_disagr | sli_agree | agree | stro_agree |
| q76 | hcw_relationship | | _ | stro_disagr | disagr | sli_disagr | sli_agree | agree | stro_agree |
| q77 | hcw_experienced | | _ | stro_disagr | disagr | sli_disagr | sli_agree | agree | stro_agree |
| q78 | hcw_likes_me | | _ | stro_disagr | disagr | sli_disagr | sli_agree | agree | stro_agree |
| q79 | hcw_distant | | _ | stro_disagr | disagr | sli_disagr | sli_agree | agree | stro_agree |
| q80 | CD4_what | text | | | | | | | |
| q81 | ARV_CD4 | | _ | up | down | | | | |
| q82 | VL_what | text | | | | | | | |
| q83 | ARV_VL | | _ | up | down | | | | |
| q84 | current_med_HIV | text | | | | | | | |
| q85 | take_ARV_bad | | _ | agree | unsure | disagr | | | |
| q86 | take_ARV_tired | | _ | agree | unsure | disagr | | | |

| | | | | | | | | |
|---|---|---|---|---|---|---|---|---|
| q87 | take_ARV_down | | _ | agree | unsure | disagr | | |
| q88 | take_ARV_taste | | _ | agree | unsure | disagr | | |
| q89 | take_ARV_good | | _ | agree | unsure | disagr | | |
| q90 | mental_interest | | _ | not_at_all | sev_da ys | half_da ys | every_d ay | refuse |
| q91 | mental_depressed | | _ | not_at_all | sev_da ys | half_da ys | every_d ay | refuse |
| q92 | mental_sleep | | _ | not_at_all | sev_da ys | half_da ys | every_d ay | refuse |
| q93 | mental_tired | | _ | not_at_all | sev_da ys | half_da ys | every_d ay | refuse |
| q94 | mental_appetite | | _ | not_at_all | sev_da ys | half_da ys | every_d ay | refuse |
| q95 | mental_me_failur e | | _ | not_at_all | sev_da ys | half_da ys | every_d ay | refuse |
| q96 | mental_concentra tion | | _ | not_at_all | sev_da ys | half_da ys | every_d ay | refuse |
| q97 | mental_slow_restl ess | | _ | not_at_all | sev_da ys | half_da ys | every_d ay | refuse |
| q98 | mental_suicide | | _ | not_at_all | sev_da ys | half_da ys | every_d ay | refuse |
| q99 | stigma_dirty | | _ | stro_disag r | disagr | agree | stro_agr ee | |
| q100 | stigma_cursed | | _ | stro_disag r | disagr | agree | stro_agr ee | |

| | | | | | | | |
|---|---|---|---|---|---|---|---|
| q101 | stigma_trust | | _ | stro_disagr | disagr | agree | stro_agree |
| q102 | stigma_similar | | _ | stro_disagr | disagr | agree | stro_agree |
| q103 | stigma_ashamed | | _ | stro_disagr | disagr | agree | stro_agree |
| q104 | stigma_not_guilty | | _ | stro_disagr | disagr | agree | stro_agree |
| q105 | stigma_weak | | _ | stro_disagr | disagr | agree | stro_agree |
| q106 | stigma_safe_children | | _ | stro_disagr | disagr | agree | stro_agree |
| q107 | stigma_restriction s | | _ | stro_disagr | disagr | agree | stro_agree |
| q108 | stigma_punishmen t | | _ | stro_disagr | disagr | agree | stro_agree |
| q109 | stigma_isolated | | _ | stro_disagr | disagr | agree | stro_agree |
| q110 | stigma_allow_wor k | | _ | stro_disagr | disagr | agree | stro_agree |
| q111 | stigma_friendship | | _ | stro_disagr | disagr | agree | stro_agree |
| q112 | stigma_family_car e | | _ | yes | no | | |
| q113 | stigma_disclosure_family | | _ | yes | no | | |
| q114 | no_sex_partner_12m | 0-50 | | | | | |
| q115 | no_sex_partner_1m | 0-10 | | | | | |

| | | |
|---|---|---|
| q116a | initial_part1 | text |
| q116b | initial_part2 | text |
| q116c | initial_part3 | text |
| q116d | initial_part4 | text |
| q116e | initial_part5 | text |
| q116f | initial_part6 | text |
| q116g | initial_part7 | text |
| q116h | initial_part8 | text |
| q117a | age_part1 | 0-100 |
| q117b | age_part2 | 0-100 |
| q117c | age_part3 | 0-100 |
| q117d | age_part4 | 0-100 |
| q117e | age_part5 | 0-100 |
| q117f | age_part6 | 0-100 |
| q117g | age_part7 | 0-100 |

| | | |
|---|---|---|
| q117h | age_part8 | 0-100 |
| q118a | sex_part1 | 0-62 |
| q118b | sex_part2 | 0-62 |
| q118c | sex_part3 | 0-62 |
| q118d | sex_part4 | 0-62 |
| q118e | sex_part5 | 0-62 |
| q118f | sex_part6 | 0-62 |
| q118g | sex_part7 | 0-62 |
| q118h | sex_part8 | 0-62 |
| q119a_y | years_part1 | 0-90 |
| q119b_y | years_part2 | 0-90 |
| q119c_y | years_part3 | 0-90 |
| q119d_y | years_part4 | 0-90 |
| q119e_y | years_part5 | 0-90 |
| q119f_y | years_part6 | 0-90 |
| q119g_y | years_part7 | 0-90 |
| q119h_y | years_part8 | 0-90 |
| q119a_m | months_part1 | 0-12 |

| | | |
|---|---|---|
| q119b_m | months_part2 | 0-12 |
| q119c_m | months_part3 | 0-12 |
| q119d_m | months_part4 | 0-12 |
| q119e_m | months_part5 | 0-12 |
| q119f_m | months_part6 | 0-12 |
| q119g_m | months_part7 | 0-12 |
| q119h_m | months_part8 | 0-12 |
| q119a_d | days_part1 | 0-31 |
| q119b_d | days_part2 | 0-31 |
| q119c_d | days_part3 | 0-31 |
| q119d_d | days_part4 | 0-31 |
| q119e_d | days_part5 | 0-31 |
| q119f_d | days_part6 | 0-31 |
| q119g_d | days_part7 | 0-31 |
| q119h_d | days_part8 | 0-31 |

| | | | | | | | | |
|---|---|---|---|---|---|---|---|---|
| q120a | condom_part1 | | _ | always | half_time | rarely | never | |
| q120b | condom_part2 | | _ | always | half_time | rarely | never | |
| q120c | condom_part3 | | _ | always | half_time | rarely | never | |
| q120d | condom_part4 | | _ | always | half_time | rarely | never | |
| q120e | condom_part5 | | _ | always | half_time | rarely | never | |
| q120f | condom_part6 | | _ | always | half_time | rarely | never | |
| q120g | condom_part7 | | _ | always | half_time | rarely | never | |
| q120h | condom_part8 | | _ | always | half_time | rarely | never | |
| q121 | sex_money | | _ | never | sometimes | often | always | |
| q122 | alcohol | | _ | never | less_m | monthly | weekly | daily |
| q123 | alcohol_six | | _ | never | less_m | monthly | weekly | daily |
| q124 | alcohol_memory | | _ | never | less_m | monthly | weekly | daily |
| q125a | use_dagga | | no | yes | | | | |
| q125b | use_benzene | | no | yes | | | | |
| q125c | use_mandrax | | no | yes | | | | |
| q125d | use_iv | | no | yes | | | | |
| q125e | use_nyaope | | no | yes | | | | |
| q125f | use_glue | | no | yes | | | | |

| | | | | |
|---|---|---|---|---|
| q125g | use_other | | no | yes |

**Phase III codebook: Participant’s understanding of adherence (and their recommendations to improve adherence to medication) (in-depth interviews)**

| Themes | Guide | Codes |
|---|---|---|
| **Participant’s life context** | Information about participant's time on ART and mentioning their age, gender, ethnicity and how long you have been on medication. Investigation on their knowledge about ART | **Participant's Life context**<br>**Age**<br>**Gender**<br>**Ethnicity and**<br>**Duration on ART**<br>**Knowledge about ART** |
| **Treatment history** | Information about the time the particpant was diagnosed with HIV, when they started on treatment, (include the time it took to start treatment). Discussing the time treatment was interrupted or stopped and reasons for interruptions | **Treatment history**<br>**HIV diagnosis**<br>**ART start information**<br>**Treatment interruptions**<br>**Adherence counselling**<br>**Reasons for starting treatment,**<br>**Duration it took to start ART (and reasons for taking longer/shorter),** |

| | | |
|---|---|---|
| | If ever stopped ART for a month or longer, reasons for restarting ARVs again? | **Restarting ART,**<br>**Health complications,**<br>**Self-motivation,**<br>**Adherence counselling,** |
| | | **Support from relative/family member/partner/friends** |
| **Current use of ART** | Experiences on taking ART medication. What makes participants take their medication. Also discussing current challenges with taking ART correctly or as prescribed | **Factors associated with ART adherence (facilitators and barriers to ART adherence).**<br>**Treatment taking behaviour,**<br>**Motivation to take treatment**<br>**Current challenges or difficulties with taking ART,**<br>**Effects of comorbidities**<br>**Distance to health facilities**<br>**Education on ART adherence** |
| | Discussing on the feeling of taking ART/ARVs. | **Regimen related factors**<br>**Effect of size of the pill(s)**<br>**Pill burden**<br>**Dosages**<br>**Frequencies**<br>**Side effects.** |
| **Individual level factors (intrapersonal)** | Discussing the participant's views on substance use (alcohol and drugs) while taking ART or replacing treatment with other non-ART medications or stopping treatment completely? | **Substance use (alcohol and drugs)**<br>**Non-ART medication**<br>**Cultural beliefs and spirituality**<br>**Effects of substance and drug abuse (towards ART adherence) and using non-ART medications, effects** |

| | | |
|---|---|---|
| | Discusing the views on the role of cultural beliefs and spirituality towards ART adherence? | **Influence of religion and cultural beliefs on adherence (both positive and negative)** |
| | Discussing the role of having employment, financial means or reliable transport to a health facility affects adherence to treatment | **Financial related/economic factors**<br>**Effects of lack of financial support**<br>**Lack of employment**<br>**Poverty** |
| | | **Lack of transportation to a health facility.** |
| **Relationships level factors (interpersonal)** | Discussing the role of treatment supporters? Or people who help take treatment better? It could be family members, partner, friend or health care worker? | **Treatment support,**<br>**Role of treatment supporters (family members, partner(s), relatives, friends and health care workers.**<br>**Trust**<br>**Confidentiality**<br>**Communication** |
| **Community-level factors** | Discussing how the community/society feel about people with HIV or on ART?<br><br>The impact of discrimination and stigma towards ART patients?<br><br>Discussig issues around disclosure of HIV status and taking ART<br><br>Discussing veiws about ART services available within the health facility (including adherence services)? | **Societal norms/stigma**<br>**Discrimination and disclosure,**<br>**Societal perceptions and views**<br>**Social norms**<br>**Poverty,**<br>**Disclosure,**<br>**Access and availability of health services within health facilities.** |

| | | |
|---|---|---|
| **Policy level factors (Macrolevel factors or healthcare policy factors)** | Discussing views on the role of ART policies and guidelines towards ART adherence? | **Guidelines and policies,**<br>**Participants have knowledge and an understanding of the role of policies and guidelines**<br>**Guidelines play a role towards ART adherence** |
| **Treatment taking reminders** | Discussing treatment taking reminders if any | **Treatment taking reminders,**<br>**treatment storage**<br>**Reasons for using the specific reminders and storage.** |
| **Recommendations for adherence strategies** | Discussing what participants think should be done to improve adherence of patients on ART? | **Recommendations for adherence strategies,** |
| **Closing** | Discussing any other issues a participant would like to raise regarding ART? | |

### Phase IV codebook: Systematic review data collection (article extraction form)

| | # | Variable / Field Name | Field Label *Field Note* | Field Attributes (Field Type, Validation, Choices, Calculations, etc.) |
|---|---|---|---|---|
| Instrument: Article extraction form | | | | |
| | 1 | record_id | Article number | text |
| | 2 | authors | Authors | text |
| | 3 | title | Title | text |
| | 4 | year_published | Year article published | text |
| | 5 | abstract | Abstract | notes |
| | 6 | journal | Journal | text |
| | 7 | objectives | Objectives | text |
| | 8 | population | Study population | text |
| | 9 | Study design | Study design | text |
| | 10 | sample_size | Sample size | text |
| | 11 | adherence_strategy1 | Adherence strategy 1 | text |
| | 12 | adherence_strategy2 | Adherence strategy 2 | text |
| | 13 | adherence_strategy3 | Adherence strategy 3 | text |
| | 14 | adherence_strategy4 | Adherence strategy 4 | text |
| | 15 | adherence_strategy5 | Adherence strategy 5 | text |
| | 16 | comparison_s | Comparison(s) | text |
| | 17 | outcome | Outcome | text |
| | 18 | comments | Comments | notes |
