## Supplementary appendix 2 for "Study protocol: Strengthening understanding of effective adherence strategies for first-line and second-line antiretroviral therapy (ART) in selected rural and urban communities in South Africa"

**Semi-structured Interview guide**

**INSTRUCTIONS**

1. This interview is intended to be an informal conversation to collect information regarding factors (facilitators and barriers) associated with adherence on first line and second line ART regimen and strategies to improve adherence to treatment
2. Each interview must be audio recorded ONLY after the informed and recording consent form have been signed.
3. There are two levels of questions:
    a. **Main questions: the questions that the investigators want to get answers to**
    b. **Probes: to assist the moderator to get greater clarity on certain issues and more information about the main question**

***If the interviewer is different from the person consenting:***

Hello, my name is ____________________________ and I am an employee at Ezintsha, a subdivision of Wits Reproductive Health and HIV Institute (Wits RHI). I would like to invite you to participate in an interview to understand factors (facilitators and barriers) associated with adherence on first line and second line regimen and strategies to improve adherence to treatment.

Please note that I simply want to hear your views, and there is no right or wrong answer; everything you say is important. Please feel free to speak openly and use any language or words that best describe your experiences and views. Your real name will not be written anywhere, which means that no one will know it was you who said something. I will like to record the interview using an audio recorder to help me remember all the information from our conversation. We will transcribe (write out) what you have said. The voice files and notes will be kept private and safe. The discussion will take about 45 – 60 minutes.

**Date of interview (DD/MM/YYYY): ______________ Client initials: ____________ Clinic name: ___________________**

**Tier or study unique #: ____________________ Viral load status (VLS or VLF) ___________________**

**Last viral load count (number, check the patient file or TIER.Net): ___________**

**Guide #________________________**

| Themes | Sub-themes | Main questions<br>***Probes***<br>*[Use probes when you need to clarify the main question or to obtain more information from the participant]* | Notes |
|---|---|---|---|
| **Participant's life context** | **Life context** | Tell me a bit about yourself (not mentioning your name but maybe your age, gender, ethnicity and how long you have been on medication/ART)?<br><br>What do you understand and know about ART?<br><br>***Probe***<br>*Probe to allow sufficient introduction from participants that will include socio-demographic information (gender, age and ethnicity) and allow participants to be free and expressive.*<br>*Probe whether participants know how long they have been on ART.*<br>*Probe on whether participants know why they take ART.* | **<u>Life context</u>** |
| **Treatment history** | **HIV diagnosis, when started ART, treatment interruptions** | Please tell me about the time you were diagnosed with HIV, when you were started on treatment, (include the time it took you to start treatment)<br>Please tell me about the time you interrupted or stopped ART (if ever) for over one month. What | **<u>Treatment history</u>** |

| | | | |
|---|---|---|---|
| | | made you stop taking your treatment or interrupt treatment?<br><br>***Probe***<br>*Probe from responses provided by participants. Probe on how participants felt after diagnosis, whether adherence counselling was provided after diagnosis, discussed starting treatment with the health care worker, reasons for starting treatment, duration it took to start ART (and reasons for taking longer/shorter),*<br><br>*Probe if participants have ever interrupted and reasons for treatment interruption.* | |
| | **Restarting ART** | If ever stopped ART for a month or longer, why did you decide to restart ARVs again? *(Skip this question if participant has never stopped treatment)*<br><br>***Probe***<br>*Probe from responses provided by participants. Probe on reasons for restarting ART (health complications, self-motivation, adherence counselling, support from relative/family member/partner/friends)* | **<u>Restarting ART</u>** |
| **Current use of ART** | **Facilitators and barriers to ART adherence** | Tell me about your experience taking your medication. What makes you take your medication? | **<u>Factors associated with ART adherence</u>** |

| | | | |
|---|---|---|---|
| | | Tell me about your current challenges with taking ART correctly or as prescribed?<br><br>***Probe***<br>*Probe from responses provided by participants. Probe about what affects their treatment taking behaviour, motivation to take treatment, current challenges or difficulties with taking ART*<br>*Probe about effects of comorbidities, distance to health facilities, education on ART adherence (leading to other individual factors below).*<br><br>*NOTE: This section efficiently introduces socio-ecological levels (below) as participants are likely to mention related factors here.* | |
| **Individual level factors (intrapersonal)** | **Regimen related factors** | Tell me what it is like to take ART/ARVs?<br><br>How do you feel about the drugs you are taking?<br><br>***Probe***<br>*Probe from responses provided by participants. Probe about effect of size of the pill(s), pill burden, dosages, frequencies and side effects.* | **<u>Regimen related factors</u>** |
| | **Effects of substance use (alcohol and drugs), non-ART medication, cultural beliefs and spirituality towards adherence to ART** | What are your views on substance use (alcohol and drugs) while taking ART or replacing treatment with other non-ART medications or stopping treatment completely?<br><br>What are your views on the role of cultural beliefs and spirituality towards ART adherence? | **<u>substance use (alcohol and drugs), non-ART medication, cultural beliefs and spirituality</u>** |

| | | | |
|---|---|---|---|
| | | ***Probe***<br>*Probe from responses provided by participants. Probe about the effects of substance and drug abuse (towards ART adherence) and using non-ART medications (could be in addition to ART or replacing ART with non-ART medications).*<br>*Probe on the effects and influence of religion and cultural beliefs on adherence (both positive and negative).* | |
| | **Financial related/economic factors** | How do you think having employment, financial means or reliable transport to a health facility affects adherence to treatment?<br><br>***Probe***<br>*Probe from responses provided by participants. Probe about the effects of lack of financial support, lack of employment, poverty, lack of transportation to a health facility towards ART adherence.* | **<u>Financial related/economic factors</u>** |
| **Relationships level factors (interpersonal)** | **Treatment support** | Tell me about what you think of treatment supporters? Or people who help you take treatment better? It could be a family member, partner, friend or health care worker?<br><br>**Probe**<br>*Probe from responses provided by participants. Probe on participants' views about the role of treatment supporters towards ART adherence. This includes family members, partner(s),* | **<u>Treatment support</u>** |

| | | | |
|---|---|---|---|
| | | *relatives, friends and health care workers. Also probe using the following relationship factors: trust, confidentiality and communication.*<br>*Probe on the participant's relationship with health care workers at the clinic? Does it have effect on adherence to medication?* | |
| **Community-level factors** | **Societal norms/stigma, discrimination and disclosure** | How do you think the community/society feel about people with HIV or on ART?<br><br>What do you think is the impact of discrimination and stigma towards ART patients?<br><br>How do you feel about talking to other people about your HIV status (disclosure) or that you are taking ART medication?<br><br>How do you feel about ART services available within the health facility (including adherence services)?<br>***Probe***<br>*Probe on how community level factors affect ART adherence (probe from participant responses).*<br>*Probe about the societal perceptions and views on people with HIV/AIDS (or people taking ART), with more focus on social norms, poverty, disclosure, HIV related discrimination and stigma, and access and availability of health services within health facilities.* | <u>**Societal norms/stigma, discrimination and disclosure**</u> |

| | | | |
|---|---|---|---|
| **Policy level factors (Macro-level factors or healthcare policy factors)** | **Guidelines and policies** | What are your views on the role of ART policies and guidelines towards ART adherence?<br><br>***Probe***<br>*Probe from participant responses.*<br>*Probe whether participants have knowledge and an understanding of the role of policies and guidelines and participants views on whether policies and guidelines play a role towards ART adherence* | **<u>Guidelines and policies</u>** |
| **Treatment taking reminders** | | Talk to me about what you use as a reminder to take your medication? Why do you use it?<br><br>***Probe***<br>*Probe from participant responses.*<br>*Probe about what makes participants remember to take tablets, treatment storage, and reasons for using the specific reminders and storage.* | **<u>Treatment taking reminders</u>** |
| **Recommendations for adherence strategies** | | In your opinion, what do you think should be done to improve adherence of patients on ART?<br><br>***Probe***<br>*Probe from participant responses.*<br>*Probe about any adherence strategies or treatment support that participants can recommend in order to take medication correctly. It could be a strategy or treatment support that is currently offered or would like to see offered.* | **<u>Recommendations for adherence strategies</u>** |

| | |
|---|---|
| **Closing** | We have come to the end of our interview, do you have any issues that you would like to raise regarding ART?<br><br>Do you have any additional comments on how ART adherence or treatment taking behaviour can be improved? |

______________________________ ______________

Signature of interviewer Date
