## Supplementary appendix 3 for "Study protocol: Strengthening understanding of effective adherence strategies for first-line and second-line antiretroviral therapy (ART) in selected rural and urban communities in South Africa"

**Consent Sheet**

Participant/Respondent/Volunteer's statement

THE INTERVIEW HAS BEEN EXPLAINED TO ME. I HAVE BEEN GIVEN A CHANCE TO ASK ANY QUESTIONS I MAY HAVE AND I AM CONTENT WITH THE ANSWERS TO ALL OF MY QUESTIONS.

I ALSO KNOW THAT:

MY RECORDS WILL BE KEPT PRIVATE AND CONFIDENTIAL.

I CAN CHOOSE NOT TO BE INTERVIEWED, NOT TO ANSWER CERTAIN QUESTIONS, OR TO STOP THE INTERVIEW AT ANY TIME.

IF I REFUSE TO BE INTERVIEWED, IT WILL NOT AFFECT MY MEDICAL CARE / ROLE AT THE CLINIC.

I UNDERSTAND THAT THE INTERVIEWS FROM APPROXIMATELY 60 PEOPLE ON FIRST-LINE AND SECOND-LINE ANTI-RETROVIRAL THERAPY WILL BE ANALYSED, INCLUDING MINE AND REPORTED ON AS FINDINGS OF THE STUDY.

______/______/______     ______________________     _________________________________

Date     Name of volunteer/participant     Signature or Mark of Volunteer

______/______/______     ______________________     _________________________________

Date     Name of witness     Signature of Witness
(If participant is illiterate)

**Interviewer's statement**

I, the undersigned, have defined and explained to the volunteer in a language that he/she understands the procedures to be followed, the risks and benefits involved, and the obligations of the interviewer.

_______________     __________________     ________________________

Date     Name of interviewer     Signature of interviewer

**Contact Details**

Principal Investigator: Mr Siphamandla Gumede
Candidate PhD
School of Clinical Medicine, University of the Witwatersrand, Johannesburg
Interdisciplinary Social Science: Public Health; Department of Interdisciplinary Social Science, Utrecht University (UU), The Netherlands


Co-Investigator/Supervisor: Dr Samanta Lalla-Edward
Head: Research Development
Ezintsha
