## Supplementary appendix 4 for "Study protocol: Strengthening understanding of effective adherence strategies for first-line and second-line antiretroviral therapy (ART) in selected rural and urban communities in South Africa"

### Participant Information Sheet

**Introduction and purpose of the study:**
Hello, my name is ______________________________ and I am a PhD student at the University of the Witwatersrand, South Africa and Utrecht University, Netherlands. I am also an employee at Ezintsha, a subdivision of Wits Reproductive Health and HIV Institute (Wits RHI). I am recruiting 60 people to participate in a study to understand effective adherence strategies for first line and second line antiretroviral therapy (ART) in eight of the healthcare facilities in the City of Johannesburg Region F and Ndlovu Medical Centre, Elandsdoorn, Limpopo. Because you are currently taking first line or second line regimen in one of these facilities, I would like to invite you to participate in an interview, to be held at a mutually agreed upon venue, date and time. The information gathered from this interview will help to understand factors (facilitators and barriers) associated with adherence on first line and second line regimen.

This information form is to help you decide if you would like to give your permission to participate. You should fully understand what is involved before you decide to take part in this study. If you have any questions, do not hesitate to ask me. **You should not agree to take part unless you are satisfied about all the procedures involved.** If you decide to take part in this study, you will be asked to sign this document to confirm that you understand the study. Should you require it you can have a copy of this form to keep.

**Procedure:**
During the interview, we will cover two types of questions – ones in which I will give you alternatives and you chose one and other questions in which you can answer with your own answer. The interview should take 45-60 minutes. I would also like to record the interview using an audio recorder to help me remember all the information from our conversation. This will only be done however with your consent. We will transcribe (write out) what you have said. Our discussion will be confidential, and your name will not be recorded with the tape recording. There will be no way of linking what you say in this interview to who you are. Only the staff from the research study, from Ezintsha will see this information. The audio recording will be retained for a minimum of two and a maximum of six years. You may still participate in the interview should you wish not to be audio recorded. In this case, I will administer the questionnaire and write down notes as I ask you the questions.

**Consent:**
Your participation in this interview process is completely voluntary (this means you, and only you, can choose whether you would like to join this study). You may refuse to answer any specific question if you feel uncomfortable with that question. You do not have to give me a reason for refusing to answer specific questions. You can also decide to stop participating at any time. If you decide that you don't want to be part of this study, there will be no negative consequences for you and it will have no impact on your accessing health care services/working within this clinic or anywhere else. There are no right or wrong answers to any of the questions. I only want to know about your experiences, views, comments, opinions and ideas.

**Confidentiality:**
All information discussed during the interview will be kept strictly confidential; at no point will your personal details be disclosed.

Your consent form will be kept separately from all other research documents. Only I, and co-investigators will have access (through Ezintsha) to the information you have provided. During data analyse, it will all be put together without any names so that when it is reported, no person who participated in the interviews can be identified. This study protocol has been approved by the Human Research Ethics Committee (Medical) of the University of the Witwatersrand, Johannesburg.

**Benefits:**
We think that you will probably benefit by participating because many people find that it is useful to discuss their experiences, opinions and provide feedback. I believe that the information you provide will help Wits RHI and Department of Health better understand and strengthen the Anti-Retroviral Therapy services provided to patients at large.

**Risks:**
There are no known risks to participants

**Cost:**
There are no costs to you to participate in the study. If you do participate, you will be provided with R150.00 as reimbursement for your travel and time.

**Rights of the Participant:**
You may find it uncomfortable to express your personal opinions in front of an interviewer. We want you to feel comfortable so please let us know if you feel uncomfortable. You don't have to answer any questions that you don't want to discuss and you can leave the stop the interview at any time, even if it is just for a break.

**Further information:**
You may contact me, or my supervisor, at any time with any question you may have regarding this study – details below:

Principal Investigator: Mr Siphamandla Gumede


Supervisor: Dr Samanta Lalla-Edward


This study has been approved by the Human Research Ethics Committee (Medical) of the University of the Witwatersrand, Johannesburg ("Committee"). A principal function of this Committee is to safeguard the rights and dignity of all human subjects who agree to participate in a research project and the integrity of the research.
If you have any concern over the way the study is being conducted, please contact the Chairperson of this Committee who is Prof Clement Penny, who may be contacted on telephone number 011 717 2301, or by e-mail on. The telephone numbers for the Committee secretariat are 011 717 2700/1234 and the e-mail addresses are

Thank you for reading this Study Information Sheet.

October 2019
