## Supplementary appendix 5 for "Study protocol: Strengthening understanding of effective adherence strategies for first-line and second-line antiretroviral therapy (ART) in selected rural and urban communities in South Africa"

### Adherence strategies and interventions for selected chronic conditions in sub-Saharan Africa: A systematic review and meta-analysis

Siphamandla B Gumede[1,2*]; John BF de Wit[2]; WD Francois Venter[1]; Samanta T Lalla-Edward[1]; Maaike Noorman[2]

1 Ezintsha, Faculty of Health Sciences, University of the Witwatersrand, Johannesburg, South Africa
2 Department of Interdisciplinary Social Science, Faculty of Social and Behavioural Sciences, Utrecht University

Systematic review conducted by Siphamandla B Gumede

Reviewer: Maaike AJ Noorman

Reviewer/Guarantor 1: Samanta T Lalla-Edward

Reviewer/Guarantor 2: Willem Daniel Francois Venter

Reviewer/Guarantor 3: John BF de Wit

**Corresponding author*** Siphamandla B Gumede

Ezintsha
University of the Witwatersrand,
32 Princess of Wales Terrace, Parktown, Johannesburg, 2193


#### 1. Background

Adherence is widely defined as patient's ability to follow a treatment plan and take medications at prescribed times (1–3). Poor adherence to treatment is a limiting factor in the successful health outcomes of numerous health conditions, including HIV, hypertension and diabetes mellitus (DM) (4–6). Globally reports have indicated that up to 50% of treatment for chronic or long term conditions are not taken as recommended by health providers (7–9). In Sub-Saharan Africa, a wide range of barriers to adherence for chronic conditions have been reported, including adverse drug reactions, competing responsibilities, frequencies of treatment intake, tolerability, cost of treatment, food insecurity, stigma, lack of human health resources and social factors (10–12)

In the efforts to address adherence to treatment for the chronic conditions; behavioural and psychological factors, education, integrated care and patient self-management interventions have been explored (12–14). This includes behavioural rehabilitation provided by health providers to patients, integration of pyscho-social support within health programmes, and patient's knowledge about the medication and their overall satisfaction with the treatment (12,15,16). Other studies have recommended telephonic counselling and text messaging or reminders (mobile health), packaging/medication boxes, home visits, drug level monitoring, consistent clinical monitoring of patients (12,17–19). Studies focusing on ART adherence have further emphasized the importance of compliance with standard treatment guidelines (monitoring and reporting of health information (data) to promote appropriate medicine use (10,20–22).

#### 2. Research question

What treatment adherence strategies and interventions for chronic conditions have been tested and implemented in sub-Saharan Africa?

#### 3. Objective

To assess and compare adherence intervention strategies for the chronic conditions of HIV, DM and hypertension which have been tested and implemented in sub-Saharan Africa.

#### 4. Inclusion criteria

a. Population: Patients with selected chronic conditions (HIV, hypertension, DM) in sub-Saharan Africa.

b. Intervention: All interventions listed/described as adherence interventions or strategies for the conditions of HIV, hypertension, DM.

c. Comparisons: Standard of care and other interventions reported on in the review

d. Outcome: The included studies should report any measurement of adherence to chronic conditions – primarily, effects on adherence behaviour and the changes in health outcomes. There is no preferred measurement for reporting; should there be adequate statistical reporting, a meta-analysis will be considered.

e. Setting: All information from sub-Saharan Africa only will be considered for the review.

f. Language: There will be no language restrictions.

g. Date: There will be no date/time restrictions.

h. Publication status: All the documented studies will be considered and included for review. This includes peer reviewed (i.e., papers, manuscripts, and abstracts).

i. Method: The study will be designed and reported according to PRISMA. PICO will be used as a search strategy approach. This study will describe reported adherence programmes and strategies. There will be a focus on behaviour change techniques used or reported changes in process outcomes of adherence programmes and methods of implementation for HIV, hypertension and /or DM.

**5. Search strategy and selection procedure**

We will search using several electronic databases. These will include PubMed/Medline, Web of Science, Google Scholar, Scopus, and CINAHL. If necessary, we will contact study authors and request more information on individual studies. Citations and bibliographies of records will be reviewed to identify additional relevant material.

The basic search terms included will be:

*"Chronic conditions"* OR *"hypertension"* OR *"high blood pressure"* OR *"blood pressure"* OR *"arterial hypertension"* OR *"mellitus diabetes type I"* OR *"mellitus diabetes type II"* OR *"Diabetes"* OR *"Sugar"* OR *"HIV"* OR *"Antiretroviral Therapy"* OR *"Antiretroviral Treatment"* OR *"ART"* OR *"ART Programs"* OR *"ART Programmes"* AND *"adherence"* OR *"compliance"* AND *"interventions"* OR *"strategies"* OR *"odds ratio"* OR *"risk ratio"* OR *"evaluation"* OR *"impact"* OR *"effectiveness"* OR *"outcome"* AND *"sub-Saharan Africa"* OR *"sub Saharan Africa"* OR *"sub-Saharan African"* OR *"sub Saharan African"* OR *"Africa" (table 1).*

The search terms will be adjusted to suit the database being searched. An inventory with the database searched, the corresponding search criteria used, the date when the searches were conducted, and the results will be maintained. The principal investigator will do all the searches and the second reviewer will run the searches separately for comparison. The strength of the body of evidence (quality of evidence), the risk of bias and magnitude of effect will be rated and assessed using Grading of Recommendations Assessment, Development and Evaluation (GRADE) (23,24).

**6. Data collection and management**

A pre-defined data sheet will be developed for data extraction. The tool will include (but not be limited to): reference (author, title), year of publication, setting or location, sample size, intervention description, participants receiving adherence (in case of comparison). The form will be piloted prior to be used for the final searches. The principal investigator will do all the data extraction. A second and third reviewers will conduct a quality control check on the extraction and assist with the full text review of the included material.

Data quality checks will be done through RedCap (a secure web platform for building and managing research databases).

#### 7. Data storage

Data will be captured and stored electronically, and password protected in the Microsoft Excel format and/or RedCap and will only be accessible to an investigator and reviewers only. RedCap access is restricted to only those users who are registered on the system.

#### 8. Analysis

All adherence interventions or strategies will be described, based on the type of intervention implemented and the setting. The different evaluations methods will then be described in detail by comparing the type of assessments and outcome measures. If appropriate, outcome measures will be reported in terms of changes in the prevalence or reduction in the relative risk. Whenever necessary, we will calculate unadjusted risk ratios (RRs) and 95% confidence intervals (CIs) from data provided and present the outcome indicator results in forest plots. Furthermore, we will perform a sensitivity analysis to measure the robustness of our results to the choice of summary statistic and calculated unadjusted risk differences. We will apply a random- effects model to calculate summary RRs and 95% Cl. To test the robustness of the findings, we will re-run the analysis using a fixed effects model. Data will be coded and analysed used STATA version 15.1.

#### 9. Publication

The corresponding author will produce the first draft manuscript which will be commented on by all co-authors. The systematic review will be submitted to a peer-reviewed journal.

#### 10. Duration

The review is expected to take twelve months from protocol development to manuscript submission.
