## Supplementary material for "Study protocol: Strengthening understanding of effective adherence strategies for first-line and second-line antiretroviral therapy (ART) in selected rural and urban communities in South Africa": FUNDING AGREEMENT CARNIEGE AND SIDA

**December 13, 2017**

### CARTA FELLOWSHIP AGREEMENT

Following your acceptance of the CARTA Fellowship award, you are expected to read and sign this agreement for the fellowship to be activated.

I. SIPHAMANDLA BONGA ZIPHOZONKE GUMEDE (Passport No. A01991519 Of (University/Institution) UNIVERSITY OF THE WITWATERSRAND P.O. Box 3, WITS, 2050, JOHANNESBURG hereby referred to as the "Fellow" accept the CARTA Fellowship award under the following conditions:

#### A. Fellowship commencement and duration

The CARTA Fellowship is for a maximum period of four years, with an official commencement date of March 5, 2018. The Fellowship terminates 48 months after commencement or upon completion of your PhD training.

#### B. CARTA Fellowship funding

The total value of the Fellowship is up to **US$100,000.00** covering among others the following costs:

1. Participation in CARTA's four-part Joint Advanced Seminars (JAS) is compulsory. JAS will be held about once every year for periods of between 20 - 35 days. The first two seminars will be held in year 1, none in year 2 and one each in years 3 and 4. The Fellowship will cover a round-trip economy ticket from Fellows' home/university base to the venue, accommodation, meals, and training materials. Visa costs and airport transfers to and from the training venue will also be covered by the Fellowship. Failure to attend the JAS without prior permission will result to termination of the fellowship.
2. Monthly stipend of US$750.00 will be paid to the Fellow during the course of the Fellowship. The stipend will be paid to the fellow on a half-yearly basis in advance, with the first payment made around the start of JAS 1.

3. Fellows can apply to any African university within the CARTA consortium for their PhD training. Fellows studying outside their home country will receive a supplementary stipend of US$3,000.00 plus one round trip ticket of up to US$1,000 each year. It is the responsibility of the Fellow studying outside their home institution to:
    - Select PhD research topics that can be effectively supervised by the expertise at the institution of their choice and
    - Meet the entry requirements of the chosen institution.
4. Provision of seed funding for research or fieldwork of up to US$ 7,000.00. This is to support data collection related to PhD research. CARTA encourages its Fellows to seek additional funding for their doctoral research and will constantly provide Fellows with information regarding other sources of funding to support doctoral research and related costs.
5. Sponsored participation in one scientific conference subject to the Fellow having a paper accepted for oral presentation. The paper should be related to the Fellow's PhD work. The maximum budget for participation in a scientific conference is set at US$ 3,000.00
6. Competitive internships at CARTA participating institutions. Internships will be approved by the CARTA based on:
    6.1. The quality of Fellow's proposal
    6.2. Fellow's progress in his/her PhD work, and
    6.3. How critical the internship will be to the Fellow's PhD research and professional development.

    The maximum provision for each internship award is US$10,500 and this can be taken at any time after JAS 2. Further details of the internships are available from the Secretariat.
7. CARTA does not cover tuition charges for Fellows registered at their home university. For those studying at another CARTA university, only tuition costs charged by the university shall be covered.
8. All promises of financial and other support to Fellows are subject to meeting the clear milestones set by CARTA, participation in JAS and inter-JAS activities, and completion of all CARTA deliverables and reporting requirements.
9. The CARTA Fellowship does not cover costs for the following:
    - Books

- General travels
- Health insurance at home institution
- Accommodation at the university

**C. CARTA Fellowship funding**

The CARTA Fellowship is awarded on condition that the Fellow accepts the following terms and conditions of the Fellowship:

1. The Fellow shall provide full details of his/her bank account to the African Population and Health Research Center (APHRC) to facilitate fund disbursement.
2. The Fellow shall submit progress reports every six months. These reports shall be duly signed by the Fellow, the supervisor(s) and the head of his/her department. At a minimum, the report shall provide details of the Fellow's progress on his/her PhD studies, inter-JAS activities, and timelines to completion. The first progress report will be submitted no later than August 31, 2018. Should there be a significant shift in project timelines, the Fellow will be expected to submit a letter of justification endorsed by the supervisor(s) together with the progress report. The progress report sent to CARTA secretariat shall be copied to the CARTA focal person at the Fellow's home institution and the supervisor. CARTA Secretariat reserves the right to request additional documentation or supporting documents. Fellows whose progress is deemed to be poor may have their Fellowship suspended or terminated.
3. The Fellow shall account for all funds received under Clauses B4, B5 and B6. The Fellow must submit periodic progress and financial reports accompanied by scanned copies of original receipts. The financial reports shall be duly signed by the Fellow, the supervisor(s) and head of the department where the fellow is registered for his/her PhD. Failure to account for the funds will lead to discontinuation of the monthly stipend.
4. The Fellow shall not be involved in any form of fraud. Any indications of fraud will lead to termination of the fellowship.
5. The Fellow shall adhere with the requirements of all CARTA policies and agree to the consequences of breaching them.

6. The Fellow may be contacted by the CARTA Secretariat at any time for purposes of follow up or to request for specific information.
7. CARTA will not offer additional funding outside the terms of this contract.
8. All presentations or publications resulting from research partially or wholly funded by CARTA must include the following acknowledgement:
"This research was supported by the Consortium for Advanced Research Training in Africa (CARTA). CARTA is jointly led by the African Population and Health Research Center and the University of the Witwatersrand and funded by the Carnegie Corporation of New York (Grant No--B 8606.R02), Sida (Grant No:54100029), the DELTAS Africa Initiative (Grant No: 107768/Z/15/Z). The DELTAS Africa Initiative is an independent funding scheme of the African Academy of Sciences (AAS)'s Alliance for Accelerating Excellence in Science in Africa (AESA) and supported by the New Partnership for Africa's Development Planning and Coordinating Agency (NEPAD Agency) with funding from the Wellcome Trust (UK) and the UK government, " The statements made and views expressed are solely the responsibility of the fellow.
9. Attendance at Joint Advanced Seminars and Assessments:
   9.1. The Fellow will attend all Joint Advanced Seminars (JAS). Where necessary, CARTA will provide travel insurance for the Fellow.
   9.2. The Fellow will complete all assignments by their stipulated deadlines. These assignments include inter-JAS activities which involve holding research training seminars for other graduate students in related disciplines at their home universities and/or contributing to sessions relevant to undergraduate or postgraduate courses.
10. The Fellow will submit at least one article from their dissertation to an international peer-reviewed journal during the duration of their CARTA Fellowship.

**D. CARTA Fellowship funding**

1. It is expected that Fellows will complete their studies in 4 years. Failure to meet set milestones may result in discontinuation of the CARTA Fellowship.
2. Fellows wishing to discontinue their participation in the CARTA program will communicate in writing to the Program Manager with a copy sent to the focal person at

their home institution. Such Fellows may be required to return all or part of their CARTA awards including the laptop and installed software licenses.

3. Fellows can only register at one of the nine African CARTA member-universities. Fellows who register outside the African member-universities will have their Fellowship terminated immediately.
4. Fellows who fail to secure registration for their PhD may have their Fellowship suspended.

### E. CARTA Fellowship funding

1. CARTA will cease payment of stipend after 48 months or upon completion of PhD, whichever comes first.
2. Where a Fellow completes PhD studies before the 48 month period, CARTA will accord the Fellow full sponsorship to participate in the remaining CARTA activities, especially the Joint Advanced Seminars.
3. Fellows who complete their PhD studies will be eligible to compete for any CARTA supported post-graduation opportunities.

CARTA
**Consortium for Advanced Research Training in Africa**
Building a vibrant multidisciplinary African Academy that is able to lead world-class research that makes a positive impact on Public and Population Health.
 Website: www.cartafrica.org

CARTA FELLOW:

I. SIPHAMANDLA BONGA Z. GUMEDE (Passport No. A01991519 ........................ of (University/Institution)... UNIVERSITY OF THE WITWATERSRAND........ University P.O. Box ...3, WITS, 2050, JOHANNESBURG have read and understood the terms and conditions of the CARTA Fellowship and I agree to adhere to them.

Signature.............................................. Date... 21/12/2017 ..................

HEAD OF DEPARTMENT:

Name....... WAP VENTER .............................. Signature..................................

Date.... 21 DEC 2017 ..........................................

DIRECTOR CARTA:

Name.................................................. Signature..................................

Date..................................................

Please scan and email a signed copy of this Agreement to no later than **January 22, 2018**. You are required to carry the original copy with you to JAS 1. Kindly note that the Fellowship and initial payment of stipends cannot start without receipt of the signed Agreement.
