## Supplementary material for "Study protocol: Strengthening understanding of effective adherence strategies for first-line and second-line antiretroviral therapy (ART) in selected rural and urban communities in South Africa": FUNDING AGREEMENT UTRECHT UNIVERSITY

**Dr. F.J. van Dijk, Managing Director**
**PO box 80140**
**3508 TC, Utrecht**
**The Netherlands**

**Telephone**
+31 30 253 6729
**E-mail**


**Date**
7 July 2020
**Subject**
Commitment of funding for joint UU-Wits PhD research

To whom it may concern,

I am writing to confirm the budget of €250,000 from the Faculty of Social and Behavioural Science, Utrecht University, committed to cover the cost of joint PhD research undertaken at Ezintsha, a subdivision of Wits Reproductive Health and HIV Institute, University of the Witwatersrand, Johannesburg.

The budget amount indicated above will be paid in multiple, periodic installments and is solely intended for the cost related to the joint UU-Wits PhD research project entitled 'Adherence to first-line and second-line antiretroviral therapy (ART) in selected rural and urban communities in South Africa: assessment of patient support needs and adherence strategies' (ART adherence study). This joint PhD research project is undertaken by Mr. Siphamandla Gumede, the PhD candidate and associate researcher at Ezintsha. The PhD candidate is supervised by Prof. dr. J.B.F de Wit, Utrecht University, Prof. Dr. W.D.F Venter, Ezintsha, Dr. S. Lalla-Edward, Ezinthsha, and Dr. A.M.J. Wensing, University Medical Centre Utrecht.

The above mentioned amount should be used as stipulated in the agreed budget, including in relation to cost of the salary of the PhD candidate; project-related costs in relation to international travel, accommodation and subsistence incurred by the PhD candidate and supervisors; domestic travel and accommodation cost for project-related data collection; cost of project-related equipment and open-access publishing; cost related to scientific, data management and administrative support required by the Ndlovu research consortium or the Ndlovu Care Group; and cost related to administrative support by WHRI or Ezintsha.

Sincerely,

Frank Jan van Dijk, PhD
Managing Director, Faculty of Social and Behavioural Sciences
Utrecht University
