## Supplementary material for "Study protocol: Strengthening understanding of effective adherence strategies for first-line and second-line antiretroviral therapy (ART) in selected rural and urban communities in South Africa": Human right research ethics clearance letter

UNIVERSITY OF THE WITWATERSRAND, JOHANNESBURG

R14/49 Mr SB Gumede

### HUMAN RESEARCH ETHICS COMMITTEE (MEDICAL)
### CLEARANCE CERTIFICATE NO. M190641

| | |
|---|---|
| **NAME:** (Principal Investigator) | Mr SB Gumede |
| **DEPARTMENT:** | School of Clinical Medicine<br>Wits Reproductive Health and HIV Research Institute |
| **PROJECT TITLE:** | Strengthening understanding of effective adherence strategies for first-line and second-line antiretroviral therapy (ART) in selected rural and urban communities in South Africa |
| **DATE CONSIDERED:** | 2019/06/28 |
| **DECISION:** | Approved unconditionally |
| **CONDITIONS:** | Evidence of study site approval presented and previous condition thus satisfied - 2020/03/19 |
| **SUPERVISOR:** | Profs J. de Wit & F Venter; Dr S Lalla-Edward |
| **APPROVED BY:** | Dr CB Penny, Chairperson, HREC (Medical) |
| **DATE OF APPROVAL:** | 2019/10/16 |

**This clearance certificate is valid for 5 years from date of approval. Extension may be applied for.**

#### DECLARATION OF INVESTIGATORS

To be completed in duplicate and **ONE COPY** returned to the Research Office Secretary on the 3rd Floor, Phillip Tobias Building, Parktown, University of the Witwatersrand, Johannesburg.
I/we fully understand the conditions under which I am/we are authorized to carry out the above-mentioned research and I/we undertake to ensure compliance with these conditions. Should any departure be contemplated, from the research protocol as approved, I/we undertake to submit details to the Committee. **I agree to submit a yearly progress report**. When a funder requires annual re-certification, the application date will be one year after the date when the study was initially reviewed. In this case, the study was initially reviewed in **June** and will therefore reports and re-certification will be due early in the month of **June** each year. Unreported changes to the application may invalidate the clearance given by the HREC (Medical).

Principal Investigator Signature ____________ Date ____________

**PLEASE QUOTE THE CLEARANCE CERTIFICATE NUMBER IN ALL ENQUIRIES**
