## Supplementary material for "Study protocol: Strengthening understanding of effective adherence strategies for first-line and second-line antiretroviral therapy (ART) in selected rural and urban communities in South Africa": Department of health letter of permission

GAUTENG PROVINCE
HEALTH
REPUBLIC OF SOUTH AFRICA

Joburg
a world class African city

### JOHANNESBURG HEALTH DISTRICT

Enquiries: Dr EM Ohaju

Hillbrow CHC: Administration Building
Cr Smith Str. & Klein Street
Private Bag X21, Johannesburg
South Africa, 2017

Wits
Human Research Ethics Committee(Medical),
University of The Witwatersrand
Johannesburg, South Africa


DRC Ref: 2019-10-005

NHRD Ref no: GP_201910_031

Dear: Mr S Gumede

**TITLE*<u>: Strengthening understanding of effective adherence strategies for first-line and second-line antiretroviral therapy (ART) in selected rural and urban communities in South Africa</u>***

Your application for research approval refers.

The District Research Committee has reviewed your application. This letter serves as an in-principle approval to access the Districts Health facilities (mentioned below) for the above project subject to following conditions:

- The facility to be visited:,**HILLBROW CHC, JEPPE CLINIC, MALVERN CLINIC, ROSETTENVILLE CLINIC, SOUTH RAND HOSPITAL, YEOVILLE CLINIC**
- This facility will be visited from **09/12/2019 to 09/12/2020**
- The research can only commence after you submit an ethics clearance certificate from a recognized institution.
- You will report to the Facility Manager before initiating the study.

| Sub District | Sub District Manager/ Area Manager | Contact No. | Cell phone |
|---|---|---|---|
| ABCEF | Ms Matlala | 011 440 1259 | 082 307 0267 |
| F(LA) | Oupa Montsioa | 011 681 8130 | 082 467 9423 |
| Southrand | Dr N Maleka | 011 681 2002 | 071 872 6649 |

**The following conditions must be observed:**

- Participants' rights and confidentiality will be maintained all the time.

- No resources (Financial, material and human resources) from the above facilities will be used for the study. Neither the District nor the facility will incur any additional cost for this study.
- The study will comply with **Publicly Financed Research and Development Act, 2008 (Act 51 of 2008) and its related Regulations**.
- You will submit a copy (electronic and hard copy) of your final report. In addition, you will submit a six-monthly progress report to the District Research Committee.
- Your supervisor and University of the Witwatersrand will ensure that these reports are being submitted timeously to the District Research Committee.
- The District must be acknowledged in all the reports/publications generated from the research and a copy of these reports/publications must be submitted to the District Research Committee.

We reserve our right to withdraw our approval, if you breach any of the conditions mentioned above.

Please feel free to contact us, if you have any further queries. On behalf of the District Research Committee, we would like to thank you for choosing our District to conduct such an important study.

Regards,

**Dr E.M Ohaju**
**Chairperson: District Research Committee**
**Johannesburg Health District**
**Date** 12/12/2019

**Mrs M.L Morewane**
**Chief Director**
**Johannesburg Health District**
**Date:** 12/12/2019

**Dr R Bismilla**
**Executive Director**
**Johannesburg Health District**
**Date:** 10/1/2020
