## Supplementary material for "Study protocol: Strengthening understanding of effective adherence strategies for first-line and second-line antiretroviral therapy (ART) in selected rural and urban communities in South Africa": Study approval from CMJA hospital (departmental approval)

**GAUTENG PROVINCE**
HEALTH
REPUBLIC OF SOUTH AFRICA

**CHARLOTTE MAXEKE JOHANNESBURG ACADEMIC HOSPITAL**

**Enquiries:**
**Ms. N. Mzila**
**Office of the Clinical Director**
**Tell: (011): 488-4812**
9th March 2020

GP_201910_031

Dear Mr. Siphamandla Gumede

**STUDY TITLE: Strengthening Understanding of Effective Adherence Strategies for First-Line Antiretroviral Therapy (ART) In Selected Rural and Urban Communities In South Africa**

Permission is granted for you to conduct the above recruitment activities as described in your request provided:

1. Charlotte Maxeke Johannesburg Academic Hospital will not anyway incur or inherit costs as result of the said study.
2. Your study shall not disrupt services at the study sites.
3. Strict confidentiality shall be observed at all times.
4. Informed consent shall be solicited from patients participating in your study.

Please liaise with the HOD and Unit Manager or sister in charge to agree on the dates and time that would suit all parties.

Kindly forward this office with the results of your study on completion of the research.

**Supported / not supported**

[signature]

**Dr. P. Africa**
**Acting Clinical Director**
**DATE:** 10/03/2020

**Approved/not approved**

[signature]

**Ms. G. Bogoshi**
**Chief Executive Officer**
**Date:** 11.03.2020
