## Supplementary material for "Study protocol: Strengthening understanding of effective adherence strategies for first-line and second-line antiretroviral therapy (ART) in selected rural and urban communities in South Africa": Study approval from South Rand hospital (departmental approval)

GAUTENG PROVINCE
HEALTH
REPUBLIC OF SOUTH AFRICA

**SOUTH RAND HOSPITAL**
1 Friars Hill Road, Rosettenville, 2149

**Enq: OFFICE OF THE CEO**
T: 011 681 2002/3
M: 072 704 3371
E:

To: Whom it may concern

**RE: PERMISSION TO CONDUCT A STUDY AT SOUTH RAND HOSPITAL (STRENGTHENING UNDERSTANDING OF EFFECTIVE ADHERENCE STRATEGIES FOR FIRST AND SECOND-LINE ANTIRETROVIAL THERAPY (ART) IN SELECTED RURAL AND URBAN COMMUNITIES IN SOUTH AFRICA**

This serves to inform you that the above study has been reviewed and permission granted. The research may be conducted at South Rand Hospital. South Rad Hospital pledges to provide the required support in terms of access.

Regards,

**Dr M Maleka**
**CEO: South Rand Hospital**
**3rd January 2020**
