## Supplementary material for "Study protocol: Strengthening understanding of effective adherence strategies for first-line and second-line antiretroviral therapy (ART) in selected rural and urban communities in South Africa": Wits Faculty of Health Science approval of study title

UNIVERSITY OF THE
WITWATERSRAND,
JOHANNESBURG

Private Bag 3 Wits, 2050

Reference: Mrs Sandra Benn


06 January 2021
Person No: 0300179A
PAG

Mr SBZ Gumede
Unit 7754
Mahogony Street
Roodekop Ext 11
1401
South Africa

Dear Mr Siphamandla Gumede

**Doctor of Philosophy: Approval of Title**

We have pleasure in advising that your proposal entitled *Strengthening understanding of effective adherence strategies for first-line and second-line antiretroviral therapy(ART) in selected rural and urban communities in South Africa* has been approved. Please note that any amendments to this title have to be endorsed by the Faculty's higher degrees committee and formally approved.

Yours sincerely

Mrs Sandra Benn
Faculty Registrar
Faculty of Health Sciences

100 1922 2022
